# Predicting COVID-19 case counts using SARS-CoV-2 genetic diversity from wastewater

**DOI:** 10.1101/2025.11.07.25339787

**Authors:** Sana Naderi, Steven G. Sutcliffe, Gavin M. Douglas, Sukriye Celikkol Aydin, Inès Levade, Judith Fafard, Lila Naouelle Salhi, Fernando Sanchez-Quete, Sarah Reiling, Ju-ling Liu, Marc-Denis Rioux, David Dreifuss, Ivan Topolsky, Niko Beerenwinkel, Selena M. Sagan, Stephanie K. Loeb, Peter Vanrolleghem, Sarah Dorner, Dominic Frigon, Jiannis Ragoussis, B. Jesse Shapiro

## Abstract

Wastewater monitoring is a promising, cost-effective method for pathogen surveillance, particularly when individual patient testing is limited. A typical approach is to estimate viral concentrations using PCR-based quantification of viral nucleic acids. This approach has been successfully used during the ongoing COVID-19 pandemic to track, and even predict, waves of infection in a community. Although simple in principle, PCR-based quantification can be noisy and susceptible to false-negatives when mutations interrupt primer binding sites. Here we compare PCR-based quantification to whole-genome sequencing of SARS-CoV-2 to predict the number of positive COVID-19 cases in a local region. To do so, we calculate a simple population genetic metric as a proxy for population expansion: the number of polymorphic nucleotide sites in the viral genome, referred to as the number of segregating sites. We show that the number of segregating sites is predictive of COVID-19 case counts from up to 5-8 days prior, in 6 out of 10 wastewater sampling locations across the province of Quebec, Canada tracked over a mean period of ∼280 days. By contrast, PCR-based viral concentrations were not significantly predictive at any sampling location in Quebec. In an independent Swiss wastewater dataset sampled over a similar duration, both segregating sites and PCR-based quantification were predictive of future case counts in 5 out of 6 locations. Together, these results suggest that PCR-based approaches may be more sensitive to region-specific differences in sampling frequency and quality, whereas sequencing-based approaches may be more robust leading indicators of infection counts.

**Importance:** Wastewater monitoring of pathogens is an increasingly important part of the epidemiological surveillance toolkit. It typically involves quantifying pathogens in wastewater by PCR, an approach that is relatively simple but can be noisy and prone to false-negatives. Here we explore an alternative approach, based on quantifying viral genetic diversity by whole-genome sequencing SARS-CoV-2 from wastewater. Using wastewater samples from Quebec and Switzerland, we show that a simple metric of genetic diversity (which tracks with population expansion and does not require any pre-defined database of viral variants) is predictive of COVID-19 case counts about a week in advance, in a majority of sampled locations. While sequencing may be more costly and labor-intensive than PCR, it provides a robust and informative tool for wastewater-based epidemiology.

## Introduction

Wastewater-based surveillance is used to track various biomarkers at the community level. Throughout the coronavirus disease 2019 (COVID-19) pandemic, wastewater surveillance of severe acute respiratory syndrome coronavirus 2 (SARS-CoV-2), the causative agent of COVID-19, gained prominence as a cost-effective complement to classic diagnostic testing of individual patients. Clinical surveillance of SARS-CoV-2 can be biased by healthcare-seeking behavior (McDonald et al., 2021) and systematic healthcare disparities (Karthikeyan et al., 2022), resulting in underdetection of asymptomatic individuals (Bertels et al., 2022). Wastewater surveillance, on the other hand, allows monitoring an arbitrarily large catchment area of people excreting viruses into sewage. It is also independent of healthcare resource availability, and largely overcomes the challenge of detecting asymptomatic infections as both symptomatic and asymptomatic individuals shed SARS-CoV-2 RNA in their feces (Park et al., 2021). As a result, SARS-CoV-2 RNA in wastewater is often a leading indicator of COVID-19 cases, allowing a prompt public health response.

The most common practice for wastewater-based surveillance of COVID-19 has been quantifying SARS-CoV-2 RNA concentrations in wastewater via reverse transcription quantitative polymerase chain reaction (RT-qPCR), with studies demonstrating strong relationships between viral concentrations and local COVID-19 incidence (Mohring et al., 2024; Wolfe et al., 2021). Viral concentration, reported as viral copy numbers, can track the overall prevalence of the virus in the community (Karthikeyan et al., 2021), but lacks the resolution to identify specific viral lineages. The accuracy of viral copy numbers can be hindered by factors including shedding heterogeneity among different populations and in-sewer processes such as RNA degradation and flow rates (Bertels et al., 2022). Quantifying viral concentration typically targets one or two genes in the SARS-CoV-2 genome. The high mutation rate of SARS-CoV-2, coupled with its global spread, has led to significant genetic diversity. Novel mutations appearing in regions of the SARS-CoV-2 genome targeted by PCR primers can cause ‘amplicon dropouts,’ ultimately reducing the sensitivity of the tests (Thakali et al., 2024). Whole-genome sequencing, although more resource intensive and time consuming, yields additional information about SARS-CoV-2 genetic diversity, including lineages and variants of concern (Baaijens et al., 2022; Karthikeyan et al., 2022). Whole-genome sequencing is typically based on hundreds of PCR amplicons spanning the whole genome, making it robust to a few individual amplicon dropouts.

Genomic sequencing of SARS-CoV-2 in wastewater has enabled the detection of prevalent lineages and their relative abundances (Karthikeyan et al., 2022). This in principle could enable early detection of variants of concern that are circulating—for both SARS-CoV-2 and other well-studied pathogens—but only allows the detection of known variants by mapping to a reference database. As a result, there remains a need for early-detection methods that are agnostic to whether a variant of concern has already been characterized. Here, we propose the number of segregating sites, a population genetic metric of diversity, as a predictor for COVID-19 incidence. The number of segregating sites is informative of very recent population expansion (Chen et al., 2015; Tajima 1989) and an excess of rare variants are observed in a population undergoing exponential population growth (Coventry et al., 2010). This metric has previously been used to accurately estimate epidemiological parameters from whole genome sequences sampled from individual patients (Park et al., 2023). We apply a similar approach to wastewater sequencing time series from locations in Quebec and Switzerland, and test the utility of the number of segregating sites to predict locally matched COVID-19 case counts.

## Results and discussion

### Overview of sampling and sequencing

We sampled and sequenced wastewater samples over a one year period from March 2022 to March 2023 from 10 wastewater sampling locations across six health regions in the province of Quebec, Canada (**Methods; Figure 1a**), including urban regions (Montreal, Laval, Quebec City, Gatineau) and more rural areas (Rimouski, Sept-Îles). The wastewater sampling locations covered high percentages of the population in the largest urban areas (Montreal: 89%; Laval: 86%), moderate percentages for smaller cities (Quebec City: 67%, Gatineau: 57%), and smaller percentages of less densely populated regions (Sept-Îles: 44%, Rimouski: 20%; **Figure 1b; Supplementary Figure S1**). After applying basic quality filters (**Methods**), we were left with 2,574 sequenced samples across the 10 wastewater sampling locations. These samples typically had >10X sequencing depth across more than 50% of the SARS-CoV-2 genome, with the exception of some samples from Quebec City (**Supplementary Figure S2**).

**Figure 1.**
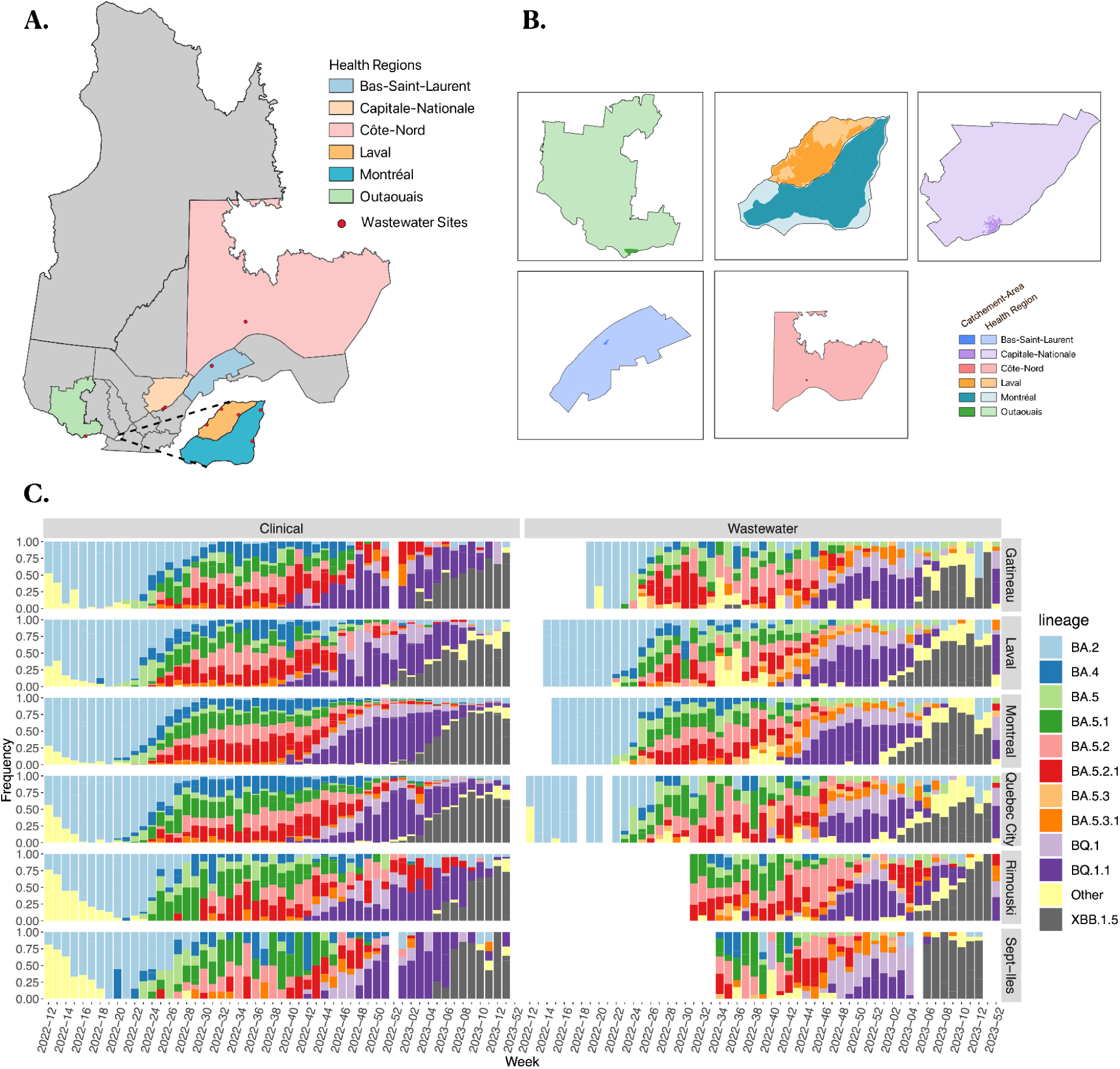
Wastewater sequencing across Quebec identifies similar dynamics of SARS-CoV-2 lineages as clinical sequencing. (a) Map of wastewater sampling locations across the province of Quebec. (b) Overlay of wastewater catchment areas (dark shaded colours) on the health regions in which positive cases are counted. (c) Relative abundances of prevalent (most abundant 11) SARS-CoV-2 lineages over a one-year time series across six health regions. Data from clinical sequences are shown on the left; data from wastewater are on the right. Lineage relative abundances (frequency) is calculated within each week of the time series.

### Both WW and clinical sequencing identify similar waves of lineage replacement

Several distinct lineages of SARS-CoV-2, including named Variants of Concern, were known to be circulating globally during our sampling period. To assess how well wastewater sequencing captured the SARS-CoV-2 lineages circulating in Quebec, we estimated their relative abundances using a computational deconvolution method (Methods; Karthikeyan et al., 2022). We then compared our wastewater-derived lineage inferences to the major lineages observed in clinical samples from Quebec hospitals over the same time period, matching them by health regions defined by the Public Health Agency of Quebec (INSPQ). We found that wastewater and clinical lineage dynamics were generally concordant, with both showing a transition from BA.2 to a mixture of BA.4 and BA.5 lineages, followed by BQ.1.1 being replaced by XBB.1.5 at the end of the sampling period (**Figure 1c**). These dynamics were similar across the six health regions in Quebec, indicating that lineage transmission was relatively homogeneous across the province, with only subtle but detectable differences (ANOSIM by health region, *R* = 0.02, *p* < 1e-4). Most of the variation occurred over time (*R* = 0.674, *p* < 1e-4; **Supplementary Figure S3**), and variation of lineages inferred in clinical versus wastewater samples was significant but minimal (*R* = 0.066, *p* < 1e-4) compared to the variation over time. This concordance of wastewater and clinical lineage calls is consistent with previous comparisons in Quebec (N’Guessan et al., 2022) and elsewhere (Baaijens et al., 2022; Crits-Christoph et al., 2021; Karthikeyan et al., 2022). Reasons for imperfect concordance include unreliable low-frequency lineage calls from wastewater – which has been previously documented and solved by filtering out lineages <5% frequency (Sutcliffe et al., 2024) – as well as potential undersampling of real diversity in clinical samples, which is more difficult to quantify.

### Predicting COVID-19 positive case counts using the number of segregating sites in SARS-CoV-2 genomes from wastewater

In population genetics, the genetic diversity found within a population can be informative of population size, and an excess of diversity is expected following population expansion. In simulated viral populations, the number of segregating sites increases with the number of infected individuals, and this metric can predict the basic reproduction number in real datasets (Park et al., 2023). Here we assess the ability of the number of segregating sites in SARS-CoV-2 genomes sampled from wastewater to predict COVID-19 prevalence in the community. To do so, we calculated the number of segregating sites, defined as the number of nucleotide sites with genetic variation in the SARS-CoV-2 genome, in each of the Quebec wastewater samples (**Methods**). The sequences considered here were filtered with more stringent quality cutoffs than in the lineage analysis described above, to ensure reliable calling of mutations within-sample. The distribution of samples retained following this step of filtering are shown in **Supplementary Figure S4**. In cities with the most dense time series (Montreal, Laval, Gatineau), sample frequency is on average 2 times a week, and in some periods is increased to daily.

To distinguish older lineage-defining mutations (that likely originated before our sampling period, outside the province of Quebec) from more recent *de novo* mutations, we counted the number of previously defined lineage-defining mutations for lineages present in the sample at 5% relative within-sample abundance or more. These lineage-defining mutations contributed less than 5% of the total number of segregating sites, on average, across the 10 sampling locations (**Supplementary Table S1**). This suggests that the vast majority of segregating sites plausibly arose by relatively recent mutation events, potentially within individual patients, and are therefore informative about recent changes in viral population size. Alternatively, some of these low-frequency mutations could have occurred less recently as parallel mutations in multiple distinct lineages. Although dozens of parallel (convergent) mutations have been documented across SARS-CoV-2 lineages (Martin et al., 2021; Wu et al., 2021), they are not numerous enough to explain the hundreds or thousands of low-frequency segregating sites observed in our data, which more likely represent recent mutations.

### Both viral copy number and the number of segregating sites correlate with COVID-19 positive case counts

To assess the relative ability of segregating sites from sequencing data and viral copy number from RT-qPCR to track infection dynamics, we calculated time-lagged correlations between each metric and COVID-19 case counts. Positive case counts and viral copy number data were obtained from Quebec Public Health (INSPQ) archives and matched to the wastewater sequences by date of collection (**Methods**). We show a representative plot of the time series for the number of segregating sites and viral copy number found in one of Montreal’s two wastewater interceptors, overlaid on the number of positive cases in the entire health region of Montreal (**Figure 2a-b**). By visual inspection, we observe that the peaks in the number of segregating sites (**Figure 2a**, dark blue line) tend to precede the peaks in the number of positive cases (violet shaded area). For viral copy number, however, the peaks in positive case counts either coincide or follow the peaks in viral copy counts (**Figure 2b**, dark red line), suggesting that although correlated, viral copy number is not necessarily predictive of positive cases.

**Figure 2.**
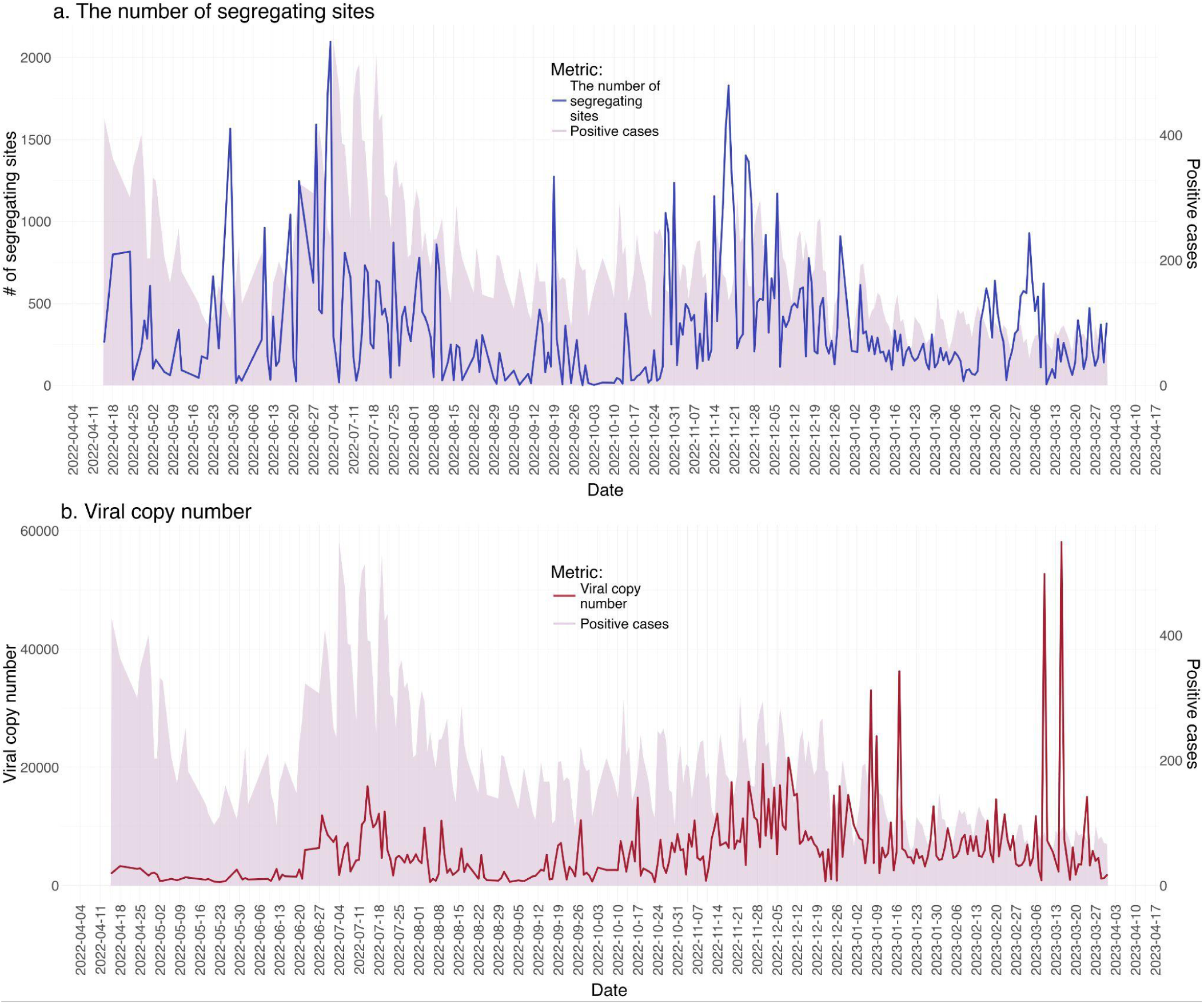
Peaks in the number of segregating sites precede peaks in viral copy number and COVID-19 case counts. Representative time series for (a) the number of segregating sites and (b) viral copy number inferred by RT-qPCR, at one of the interceptors in Montreal between April 2022 and March 2023, overlaid on the number of positive cases in this period. The violet shaded area represents the number of positive cases in both panels, and the number of segregating sites and viral copy number are plotted in blue and red, respectively. The unit of viral copy number data, are the number of gene copies of SARS-CoV-2 in billions, per 100,000 people, determined by RT-qPCR of primers targeting the N gene.

To investigate the generality of this example, we calculated lagged Spearman correlations between the number of segregating sites and positive case counts at different time lags for each wastewater sampling location. The direction of shifts in time series were defined such that correlation at positive time lags would imply that earlier changes in the number of segregating sites correlate with later changes in the number of positive case counts, and vice versa (**Methods;** schematic description in **Supplementary Figure S5**). We counted the number of wastewater sampling locations with a significant positive correlation between the number of segregating sites and positive case counts, and established a null distribution of expected correlations by way of a permutation test (**Methods**). We then conducted the same test with viral copy number as the predictor of positive case counts.

For both the number of segregating sites and viral copy number, the overall signal of correlation is stronger than expected by chance, when compared with permuted data (**Figure 3**). We observe, however, that the overall strength of positive correlation between the number of segregating sites and positive case counts, is stronger than that of viral copy number (**Figure 3**; **Supplementary Figures S6 and S7**). This is especially relevant in positive lags, where the direction of the shifts in time series points towards a predictive relationship. For lags 0 to 7, a significant, positive correlation with the number of segregating sites in a median of 7 wastewater sampling locations is observed (**Supplementary Figure S6**), indicating that higher positive case counts are associated with higher numbers of segregating sites from up to 7 time points *prior*. For viral copy number, a median of 3 wastewater sampling locations exhibit a significant positive correlation with the positive case counts between lags 0-7 (**Supplementary Figure S7**). A linear mixed model further confirms that lagged Spearman correlations are stronger for segregating sites than they are for viral copy number. We fitted the model to Spearman’s correlation estimates from lags -7 to 7, across 10 wastewater sampling locations. The fixed effect in this model was the predictor used in the Spearman’s test, and wastewater sampling location ID was a random effect. According to this model, Spearman’s correlation estimates tend to be 4.86% smaller when the correlation between viral copy number and positive case counts is tested, compared to the case where the correlation between the number of segregating sites and positive case counts is tested. This further highlights a stronger monotonic relationship between positive case counts and the number of segregating sites.

**Figure 3.**
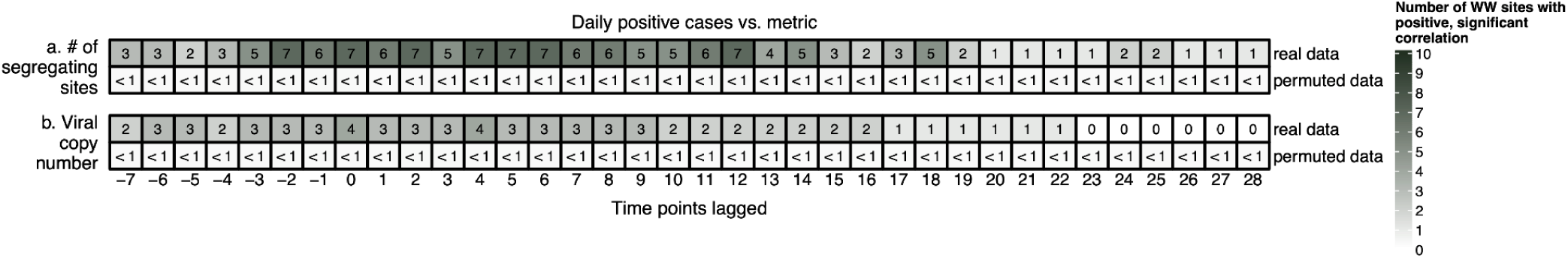
Lagged Spearman’s correlation of the number of segregating sites or viral copy number with the number of COVID-19 positive cases in Quebec. For each panel, the top row shows the number of sampling sites at the entry of wastewater sampling locations in Quebec in which a significant, positive Spearman’s correlation between the number of segregating sites (a) and RT-qPCR-derived viral copy number (b) was observed. The bottom row for each panel shows the proportion of “false-positive” tests obtained on permuted data. Lags -7 to +28 are shown, and the direction of the lags are schematically described in **Supplementary Figure S1**. The full grids for lags -28 to +28, stratified by wastewater sampling location are shown in **Supplementary Figures S6 and S7**.

Insufficient or highly variable genomic coverage is known to challenge wastewater-based surveillance of pathogens (Magbor et al., 2025). We therefore sought to determine if the observed correlation between the number of segregating sites and positive case counts is affected by the variability of coverage. To do so, we scaled the number of segregating sites to the number of genomic positions covered by a minimum of 50 reads, yielding the number of segregating sites per sufficiently covered position of the genome. Repeating the Spearman’s correlation analysis described above, we observed a generally similar pattern of significant, positive correlation, in up to 6 out of 10 wastewater sampling locations (**Supplementary Figure S8**). This analysis shows that our conclusions are robust to variable sequencing coverage across the SARS-CoV-2 genome.

### The number of segregating sites is predictive of positive cases in more sampling locations than viral copy number

Spearman’s correlations are informative of whether values of each metric (segregating sites or viral copy number) are significantly associated with the number of positive cases but they do not assess predictive value. To assess the utility of each metric in predicting future values of positive COVID-19 case counts, we applied the Granger causality test (Granger, 1969). The Granger test compares the error variance of predictions of the outcome variable (number of positive case counts) made under two scenarios (Tsay, 2013): (1) Future values of positive case counts are predicted using current and past values of positive case counts; (2) Future values of positive case counts are predicted using current and past values of an exogenous metric (in our case, either the number of segregating sites, or viral copy number) in addition to positive case counts. If the error variance is significantly reduced from scenario (1) to scenario (2), under the Granger framework, the exogenous metric is thought to *cause* the outcome (Tsay, 2013). For the purpose of our analysis, we are only interested in the predictive implications of the Granger test, and not a strict conclusion of causality. Therefore, in the presence of a significant Granger test, we conclude that the exogenous metric is useful in forecasting the number of positive cases, as incorporating information from this metric improves our predictions.

We applied the Granger test to time series from each sampled location, on orders of 1 to 28, the order being the number of time points in the past included in predictions. We found that the number of segregating sites is often predictive of positive case counts, while viral copy number is not a significant predictor (**Figure 4**). Including 5, 6, and 8 prior time points of the number of segregating sites significantly improved predictions of positive case counts in 6 out of 10 wastewater sampling locations (**Figure 4a, top row**). For viral copy number, the error variance of predictions was never significantly improved at any of the wastewater sampling locations (**Figure 4b, top row**). Similar to the Spearman’s correlation analysis, we established a null distribution of no predictivity by performing a permutation test (**Methods**). We observe a stronger predictive signal for the number of segregating sites than the corresponding null distribution (**Figure 4a**), while for viral copy number, the signal of the Granger test is weaker than the established null distribution (**Figure 4b**). To test the robustness of these results to variable sequence coverage, we scaled the number of segregating sites per sufficiently covered position of the genome, and repeated the Granger tests. We observed qualitatively similar predictive relationships after controlling for the variability of coverage across samples: in 5 out of 10 wastewater sampling locations, the number of segregating sites per sufficiently covered genomic position significantly improved predictions of positive cases (**Supplementary Figure S9**).

**Figure 4.**
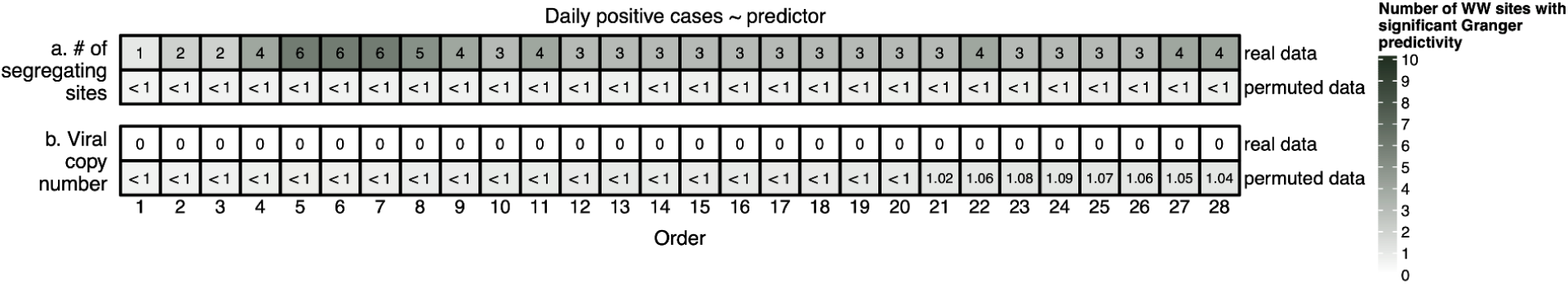
Granger test to predict COVID-19 positive case counts in Quebec. For each panel, the top row shows the number of wastewater sampling locations in Quebec in which a significant Granger relationship exists between the number of segregating sites (a) and viral copy number (b). The bottom row for each panel shows the proportion of “false-positive” tests obtained on permuted data.

### SARS-CoV-2 lineage diversity is not predictive of case counts

As we observed the co-circulation of multiple lineages, especially sub-lineages of Omicron in the studied period (**Figure 1c)**, we sought to determine if lineage diversity metrics are also informative of COVID-19 case counts. We calculated two metrics of lineage diversity; lineage richness and the Shannon diversity index (**Methods**). Lineage richness and Shannon diversity for one of the wastewater sampling locations in Montreal are shown in **Supplementary Figure S10**. We performed the Granger test on this data and found that neither metric significantly predicts changes in positive case counts (**Supplementary Figure S11**). This is not surprising, as the introduction of a more transmissible variant of concern might be predictive of increased positive case counts, but such introduction events are not well captured by these lineage diversity metrics. Instead, our results suggest that the number of segregating sites, most of which do not tag major variants of concern (**Supplementary Table S1**) but rather scale with a growing local viral population, is a better predictor of future positive case counts.

### Validating the performance of the number of segregating sites using wastewater sequence data from Switzerland

Having determined the utility of the number of segregating sites in predicting case counts in wastewater data from Quebec, we sought to validate these results in an independent dataset. Our aim was to determine whether the predictive relationship observed between the number of segregating sites and case counts would be generalizable to other settings. We obtained wastewater sequence data from the wastewater surveillance program in Switzerland (Jahn et al., 2022). Case counts and viral copy number data were obtained from the publicly available data dashboard of the Swiss Federal Office of Public Health (FOPH; **Methods**). The 6 wastewater sampling locations included in this analysis are Lugano, Werdhölzli (Zurich), Aire (Geneva), Chur, Altenrhein, and Laupen (Bern). After matching with available sequence data by date and sampling location, and excluding any periods with long interruptions in sample collection, the resulting time series covered the period from early February 2022 through late December 2022 (**Supplementary Figure S12**). The sampling frequency during this period is dense, with samples collected almost daily, except for two brief interruptions in May and September (**Supplementary Figure S12**).

Similar to the analysis from Quebec, both the number of segregating sites and viral copy number were correlated with case counts in at least 5 out of the 6 Swiss wastewater sampling locations (**Figure 5**). Unlike in Quebec, the overall signal of correlation is more consistent for viral copy number, as all 6 wastewater sampling locations show a significant correlation with case counts, on lags -7 to 17 (**Figure 5**).

**Figure 5.**
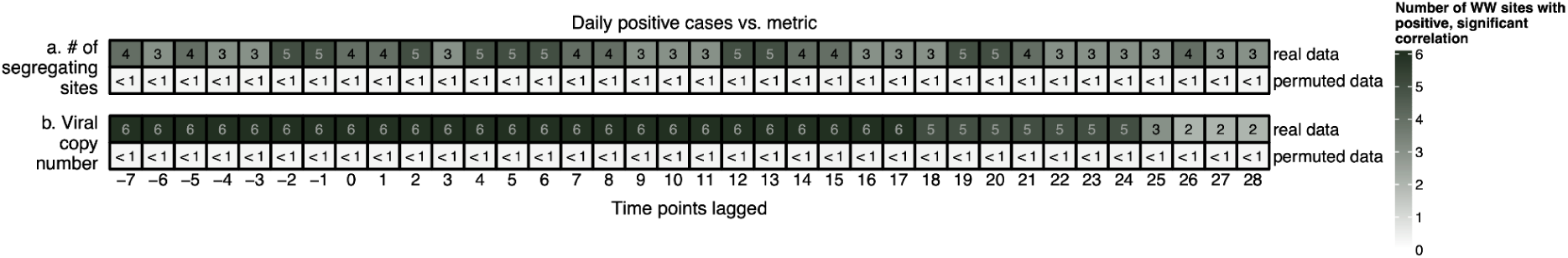
Lagged Spearman’s correlation of metrics with the number of COVID-19 positive cases in Switzerland. In each panel, the top row shows the number of wastewater sampling locations in Switzerland in which a significant, positive Spearman’s correlation between the number of segregating sites (a) and viral copy number (b) was observed. The bottom row on each panel shows the proportion of “false-positive” tests obtained on permuted data. Lags -7 to +28 are shown, and the direction of the lags are schematically described in **Supplementary Figure S1**.

The Granger test revealed a significant improvement in case count predictions that included the number of segregating sites, in 5 out 6 Swiss wastewater sampling locations (**Figure 6a**). This significant relationship is observed when 3 to 6 time points in the past were used to make predictions (**Figure 6a**). Unlike in Quebec where the inclusion of viral copy number never significantly improved predictions (**Figure 4b**), in up to 6 of the Swiss wastewater sampling locations, viral copy number is significantly informative of case counts, if up to 6 time points in the past are used in the forecast. (**Figure 6b**).

**Figure 6.**
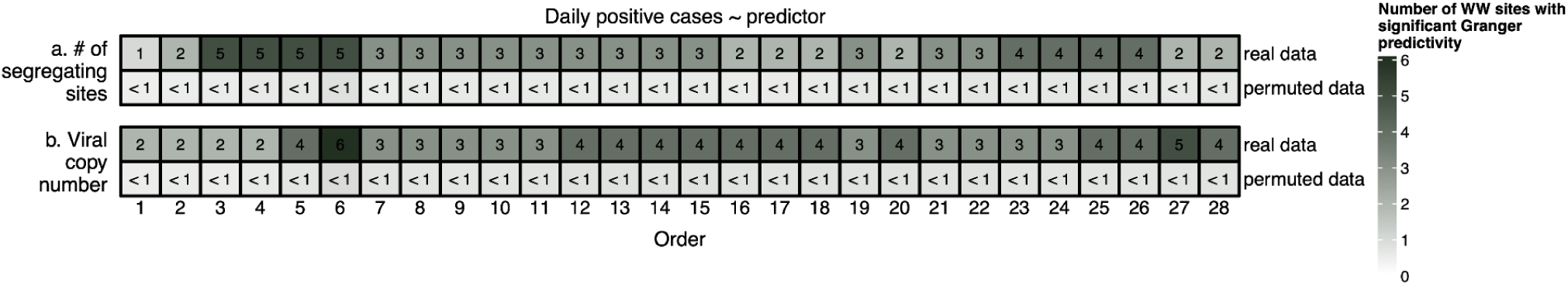
Granger test to predict COVID-19 positive case counts in Switzerland. For each panel, the top row shows the number of wastewater sampling locations in Switzerland in which a significant Granger relationship exists between the number of segregating sites (a) and viral copy number (b). The bottom row for each panel shows the proportion of “false-positive” tests obtained on permuted data.

Taken together, the analysis of two datasets demonstrate that, while the predictive utility of viral copy number as a predictor of case counts is variable across settings, the number of segregating sites consistently improves predictions, despite potential variation in wastewater systems, sampling, and sequencing strategies (**Methods**). These findings support the number of segregating sites as a robust predictor of case counts. The increased predictive utility of viral copy numbers in Swiss wastewater could be attributed to the denser temporal sampling in this dataset, or differences in wastewater retention time, dilution, flow rates, temperature, or any other differences that could affect RNA stability. Our conclusions confirm previous reports of viral copy numbers predicting or correlating with case counts (Mohring et al., 2024; Wolfe et al., 2021), but highlight that viral copy numbers may be less robust to noise, thus the lack of consistency across the two datasets presented here.

### Caveats and Conclusions

Our analysis revealed that viral copy number counts correlate with the number of cases and can be predictive of future case counts in Swiss, but not Quebec wastewater. In contrast, the number of segregating sites was a significant predictor of positive case counts both in Quebec and Switzerland, despite differences in precisely how wastewater was sampled and sequenced.

These results align with our hypothesis that viral copy numbers would be a noisier source of information compared to the number of segregating sites. Studies have discussed heterogeneity in viral shedding, both in terms of duration and quantity (Chen et al., 2024; Wölfel et al., 2020). Fecal shedding can persist for up to two weeks after recovery (Bertels et al., 2022), further complicating predictions. The number of segregating sites, however, might be a more reliable marker of community spread. This metric has previously been used to accurately estimate the basic reproduction number (Park et al., 2023). Moreover, the within-host diversity of SARS-CoV-2 is typically limited, with a median of 1-2 iSNVs within each infected person (Lythgoe et al., 2021; Naderi et al., 2024; Valesano et al., 2021). These iSNVs are rarely shared between individuals and most are unique to individual samples (Braun et al., 2021; Lythgoe et al., 2021), indicating that the number of iSNVs can provide a marker of individual infections. Using the number of segregating sites further alleviates the noise of extended shedding post recovery, as once active replication of the virus ends, no more iSNVs should be produced.

The variable sampling frequency in the wastewater data from Quebec affects our interpretation of the results in two ways. First, the lags and orders at which the Spearman’s and Granger tests are significant can not be interpreted at exactly 1-day units. We interpret these time lags and orders with caution, as the sampling frequency varied over time and across different wastewater sampling locations, and the actual time span between two time points is not always one day. This means that a significant Granger test, for instance at lag 5, does not equate to a precise 5-day forecast and could be up to two weeks, depending on the sampling location in Quebec. In Switzerland, where samples were always taken daily, we observe the most significant forecasts in the 3-6 day range, with less consistent forecasts up to 15 days. Together, this suggests that the number of segregating sites can predict case counts on the order of one week in advance. Second, while the number of segregating sites could, in fact, be predictive of case counts in more than six wastewater sites, this signal is potentially obscured by insufficient sampling at certain locations. After applying quality control filters, the wastewater sampling locations in Montreal, Laval, and Gatineau have the most densely sampled time series, and wastewater sampling locations in Quebec City are the most sparse. Previous studies have concluded that for the detection of a significant trendline slope to be possible, a minimum sampling frequency of twice a week is required (Feng et al., 2021). Therefore, we are generally more confident in results obtained for wastewater sampling locations in Montreal, Laval, and Gatineau, which also tend to be the locations in which our predictive tests are significant.

We focused our analyses on the absolute number of positive COVID-19 cases as the response variable that we wish to predict. Positivity rate–the number of positive cases divided by total tests taken on a given day–could alternatively be used to track the prevalence of infection. The choice between absolute case counts or positivity rate is ultimately contingent on determining the measure that more accurately represents the true disease load in the community. Throughout the pandemic, clinical testing (and specifically PCR-based testing of individual patients) was regulated by policies determining the eligibility of individuals being tested. These policies evolved over time, and eligibility criteria often involved factors such as clinical symptoms, demographics, occupation, and non-pharmaceutical interventions. We chose to focus our analysis on absolute case counts as opposed to positivity rate, reasoning that dividing by the number of tests adds a source of variability that is not necessarily informative of true infection spread. It has therefore been suggested that positivity rate may be biased, and the number of tests taken should be adjusted for the number of tests available (Schill et al., 2023). We further reason that by testing all time series for stationarity and correcting non-stationarity as needed, we correct for changes in baseline testing levels to some extent (**Methods**).

Going forward, it is clear that sequencing-based wastewater surveillance provides a rich source of information. While more cost-effective than individual-based sequencing, it is still more costly than less information-rich PCR-based approaches. Here we show an added benefit of sequencing-based surveillance: its ability to robustly predict case counts across two distinct time series. We hope that future work will continue to test and develop our approach, and apply it to pathogens beyond SARS-CoV-2.

## Methods

### Sample collection

We collected wastewater samples from 10 locations across the province of Quebec between March 2022 and March 2023. Raw wastewater samples were collected as previously described (N’Guessan et al., 2022, Nguyen Thanh et al., 2025). Briefly, daily composite samples were collected using an auto-sampler in flow proportional mode over a period of 24 hours. The samples from Quebec City (2 stations), Montreal (2 separate raw wastewater inputs from north and south interceptor sewers), Laval (3 stations), Gatineau (1 station), Rimouski (1 station) and Sept-Îles (1 station) were transported to the laboratory in cold chain (at 4°C). A map of wastewater sampling locations was generated using QGIS (QGIS Development Team, 3.32.3-Lima).

### Sample processing and sequencing

SARS-CoV-2 whole genome amplicon sequencing was performed using the ARTIC V4.1 protocol and sequenced on an Illumina NovaSeq 6000. Full details of these procedures, including specific modifications and quality control measures, are outlined in the Supplementary Methods.

### Sequence processing and lineage calling

Sequenced reads were analyzed using a wastewater analysis pipeline that initially removed low-quality and human-mapping reads. The remaining reads were aligned to the SARS-CoV-2 reference genome, cleaned, and then samples were filtered using permissive cutoffs to ensure sufficient data quality, retaining 2,574 samples. Finally, variant calling, lineage prediction, and relative abundance estimation, was done, with low-abundance lineages binned with their parent lineage. Details regarding the specific filtering cutoffs and subsequent lineage-based analysis are outlined in the Supplementary Methods.

### Principal Coordinates Analysis (PCoA) and Analysis of Similarity (ANOSIM)

Samples were temporally grouped into four categories—Early, Early-Middle, Late-Middle, and Late—based on 16-week sampling intervals. Lineage abundance data were Hellinger-transformed, and Bray-Curtis dissimilarities were computed using the *vegan* package (Oksanen et al., 2013) in R (R Core team, 2024). PCoA was then conducted using the ape package, applying Cailliez correction to account for negative eigenvalues. ANOSIM was performed in R (R Core team, 2024) using the *anosim()* function in the *vegan* package (Oksanen et al., 2013).

### Clinical sequence data

SARS-CoV-2 genome sequences taken from individual nasopharyngeal swabs over the same sampling period as our wastewater sampling were analyzed for comparison. Sequences were obtained from the Québec SARS-CoV-2 surveillance program led by the Laboratoire de Santé Publique du Québec (LSPQ). The accession IDs associated with the clinical sequences used in this analysis are listed in **Supplementary Table S2**.

### Variant calling and the calculation of the number of segregating sites

For all time series analyses described, we narrowed down the samples discussed above to a higher quality subset of sequences to ensure more reliable SNV calling. The quality control criteria used were as follows: a minimum of 100,000 reads, less than 20% human reads, average depth of at least 20x, and a minimum breadth of 10% at 1x, with a minimum of one percent of reads mapping to SARS-CoV-2. To count the number of segregating sites, the variant files generated by Freyja were used. These files were pre-processed using a custom script (https://github.com/nf-core/viralrecon). For every sample, mutations called at a genomic position with less than 50x depth of coverage were discarded. Then, mutations were labeled as ‘acceptable’ or ‘not acceptable’, where an acceptable mutation was defined as one for which the minor allele frequency is above 1.5% or is supported by at least 25 reads. The number of segregating sites was defined as the number of genomic positions with at least one acceptable mutation.

### Richness and Shannon diversity

We used the demixing algorithm implemented in Freyja (Karthikeyan et al., 2022) to infer mixtures of SARS-CoV-2 lineages assigned based on the Pango nomenclature, and their relative abundances in each sample, with richness defined as the number of unique lineages detected in each individual. We used a 5% relative abundance cutoff to filter false lineage calls, as previously recommended (Sutcliffe et al., 2024). 45 samples only had lineage calls with abundances below 5%, and as a result, for analyses including richness and Shannon diversity, were removed from for this analysis. For consistency across all time series analysis, the same sequence quality cutoffs as described for the number of segregating sites were used (a minimum of 100,000 reads, less than 20% human reads, average depth of at least 20x, and a minimum breadth of 10% at 1x, with a minimum of one percent of reads mapping to SARS-CoV-2).

We calculated the Shannon diversity index, H, as follows:

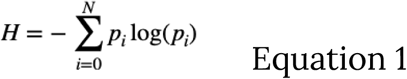

Where *p_i_* is the relative abundance of Pango lineage *i* within a sample, and the sum is taken over all *N* lineages found in a sample.

### Viral copy number data

Viral copy number data were downloaded from publicly available INSPQ archives (https://www.inspq.qc.ca/covid-19/donnees/archives) for all 10 wastewater sampling locations. As described in the Quebec Public Health (INSPQ) dashboard (https://www.inspq.qc.ca/covid-19/donnees/eaux-usees) the unit of viral copy number data, are the number of gene copies of SARS-CoV-2 in billions, per 100,000 people. The quantity of SARS-CoV-2 in wastewater is obtained by multiplying the number of copies of SARS-CoV-2 N1 gene measured per milliliter of wastewater, by the average daily flow rate in the wastewater network. The daily quantities of SARS-CoV-2 are then divided by the population served by each station, yielding per capita counts. The viral loads per capita are then multiplied by 100,000 to be reported on a basis of 100,000 people (https://www.inspq.qc.ca/covid-19/donnees/eaux-usees, under methodological details).

### Case count data

The number of daily positive case counts and daily positivity rates (the number of positive cases per the number of total tests taken on a given day) for all six health regions were obtained from the INSPQ archives (https://www.inspq.qc.ca/en/node/21994).

### Data matching and time series

The time series for the number of segregating sites, viral copy number, positive case counts, and positivity rates at every wastewater sampling location were matched and ordered by date. For health regions with more than one wastewater sampling location, the number of positive case counts and positivity rates in the entire health region was matched to the sequence data for that sampling location. The number of segregating sites and viral copy number are referred to as the ‘predictor time series’, and the number of positive case counts and positivity rates are referred to as the ‘outcome time series’ throughout.

Samples with no lineage calls with a relative abundance of 5% or more, were only dropped in analyses including richness and Shannon diversity, and were retained for the analysis of the number of segregating sites.

### Spearman’s correlation test

The Spearman’s test was applied with the *cor.test()* function in the *Stats* package in R (R Core Team., 2023). The ‘method’ argument was set to “spearman’’ and the ‘exact’ argument was set to “FALSE”. We performed lagged Spearman’s correlation tests on the time series for each of the ten wastewater sampling locations separately.

To test correlations with the number of positive case counts, two sets of tests were performed for each sampling location, one testing for lagged correlations between viral copy number (predictor) and positive case counts (outcome), and another testing for lagged correlations between the number of segregating sites (predictor) and positive case counts (outcome). In each of the tests between the pair of time series, a positive lag *k* was defined as when the predictor time series was shifted forward, such that earlier values of the predictor time series were paired with later values of the outcome by *k* lags (schematic representation in **Supplementary Figure S5**). A negative lag *k* was defined as when the predictor time series was shifted backwards, such that later values of the predictor time series were paired with earlier values of the outcome by *k* lags (schematic representation in **Supplementary Figure S5**). The Spearman’s correlation was calculated at lags ranging from -28 time points to +28 (since predictions up to a month in advance are considered feasible and useful) and for each lag, the number of wastewater sampling locations with a significant positive correlation were recorded. We ran the same workflow to compute correlations between the two predictors and positivity rate as well.

Significance was tested at a 0.05 level, using the raw p-values. Correction for multiple testing was not performed as a null distribution was generated using a permutation test (see below).

### Spearman’s correlation permutations

For each wastewater sampling location, a permutation test was done on each of the predictor-outcome combinations. For each combination, the predictor was shuffled 500 times at every wastewater sampling location, and Spearman’s correlation was tested for on the shuffled predictor and their corresponding unshuffled outcome, on lags ranging from -28 to +28 time points. The fraction of shuffled Spearman’s tests with a significant positive correlation were counted, and scaled to be represented out of 10 to be comparable with the results from the unshuffled data.

### Stationarity and the Granger test

All time series were tested for stationarity as a prerequisite for the Granger test. Stationarity was tested for using the *adf.test()* function in the *tseries* package in R (v0.10-54, Trapletti and Hornik, 1999; R Core Team, 2025), and evaluated at a 0.05 level of significance. Only non-stationary time series were differentiated, and all non-stationary time series became stationary after one degree of differentiation. The Granger test was performed using the *grangertest()* function in the *lmtest* package in R (v0.9-40, Hothorn et al., 2022; R Core Team, 2025). The Granger test was performed on all orders ranging from 1 to 28 time points. For any given predictor-outcome combination, the number of wastewater sampling locations with a significant Granger test were counted. Significance was tested at a 0.05 level, using the raw p-values. Correction for multiple testing was not performed as a null distribution was generated using a permutation test (see below).

### Granger test permutations

For each wastewater sampling location, a permutation test was done on each of the predictor-outcome combinations. For each combination, the predictor was shuffled 500 times, and the Granger test was run on the shuffled predictor and their corresponding unshuffled outcome, on orders ranging from 1 to 28 time points. The fraction of significant tests was counted, and scaled to be represented out of 10 for comparison with the results from the unshuffled data. Significance was tested at a 0.05 level.

### Swiss wastewater

We used sequencing data from the Swiss wastewater monitoring program, as previously described (Dreifuss et al., 2025; Jahn et al., 2022; John et al., 2024). The procedures used in the Swiss wastewater surveillance program to collect, sequence, and process raw wastewater sequence files is outlined in the supplementary methods section. The analysis done for our study to calculate the number of segregating sites from the available Swiss data is also outlined in the Supplementary Methods.

## Code availability

All scripts used for the preprocessing of raw sequence files and lineage analysis are available on GitHub: https://github.com/sgsutcliffe/Wastewater_Analysis_Pipeline. All scripts used for time series analysis are available on GitHub: https://github.com/Saannah/Quebecww.

## Data availability

All short-read sequencing data from Quebec wastewater are available in NCBI under BioProject PRJNA1358278. All accession IDs for the raw wastewater sequence files from the Swiss dataset are listed in Supplementary Table S3.

## Data Availability

All sequence data produced as part of this study are publicly available on the NCBI Short Read Archive.

## Acknowledgements.

We are grateful to Guillaume Bourque and José Héctor Gálvez for their contributions to the data processing stages of the raw wastewater sequence data.

