## Supplementary figures and images for "Predicting COVID-19 case counts using SARS-CoV-2 genetic diversity from wastewater"

### supp_figure_S1.pdf

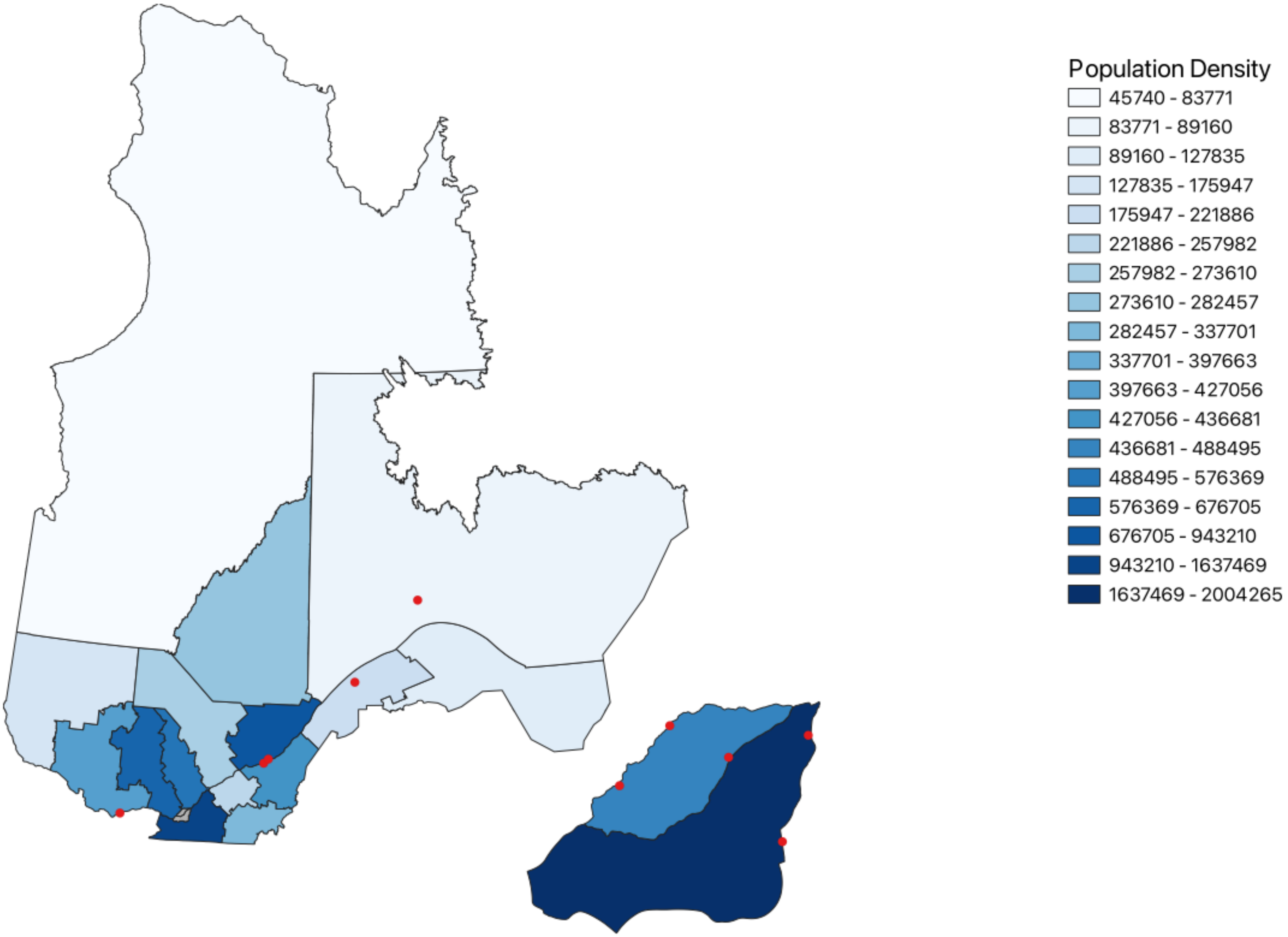

### supp_figure_S2.pdf

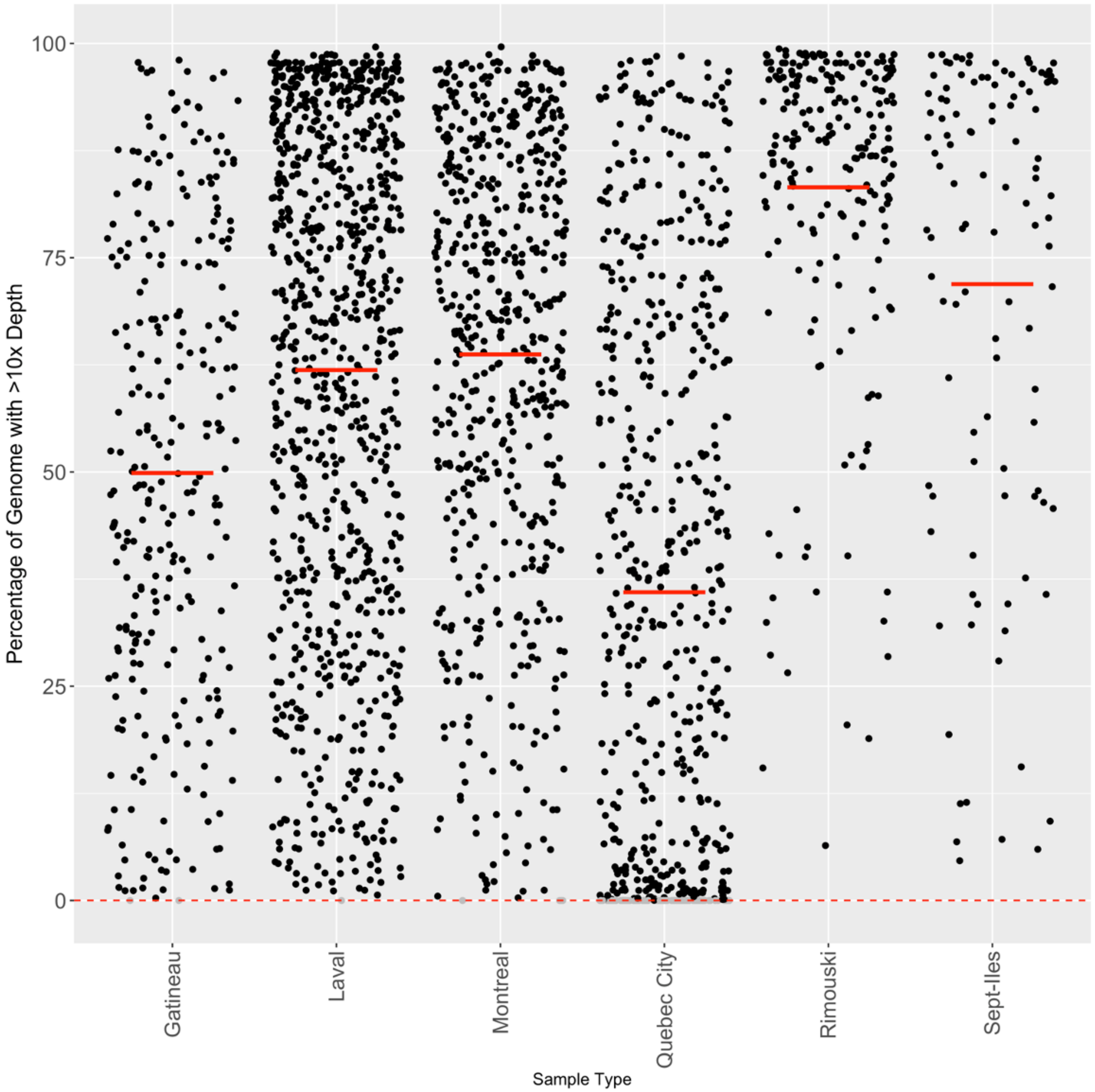

### supp_figure_S3.pdf

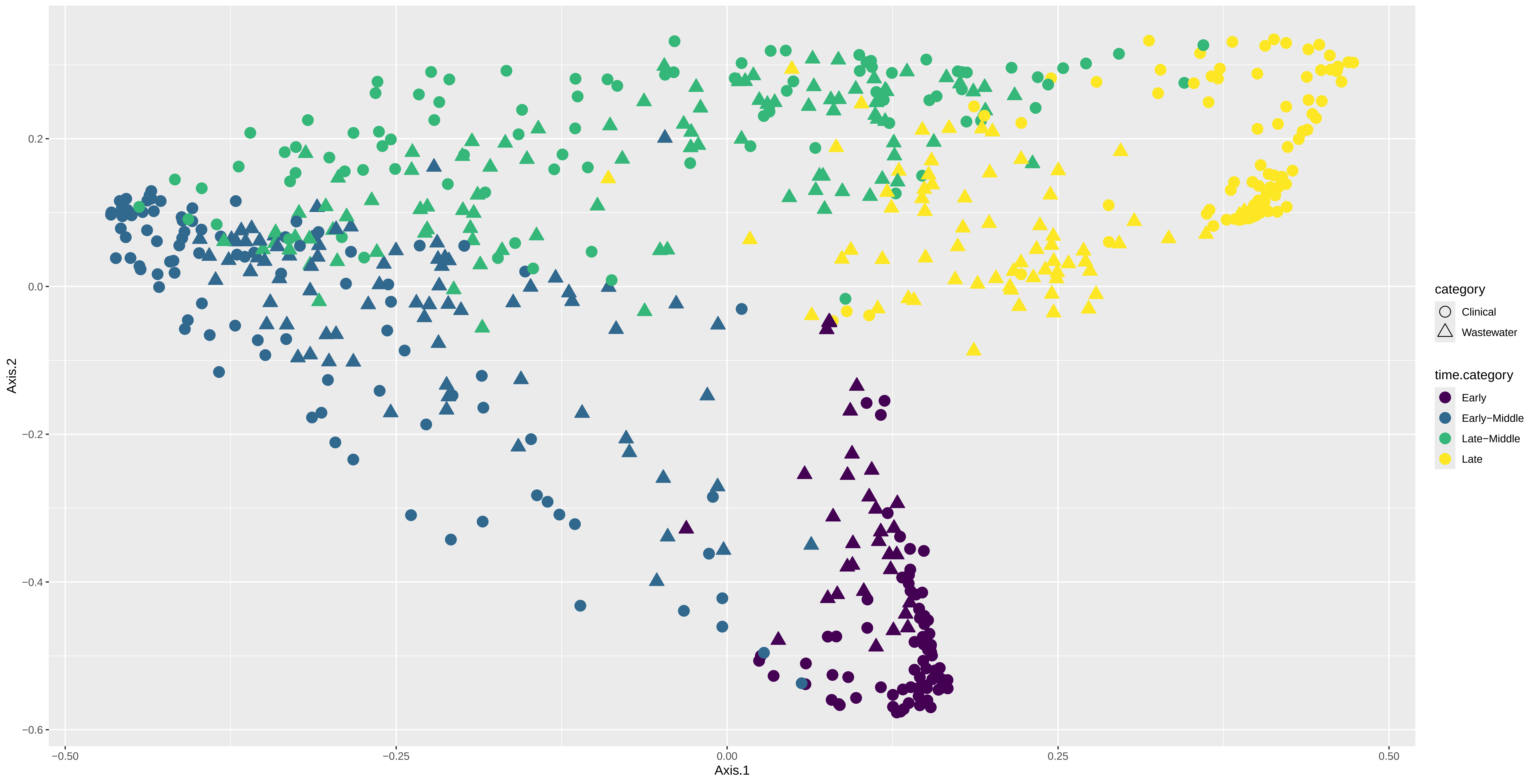

### supp_figure_S4.pdf

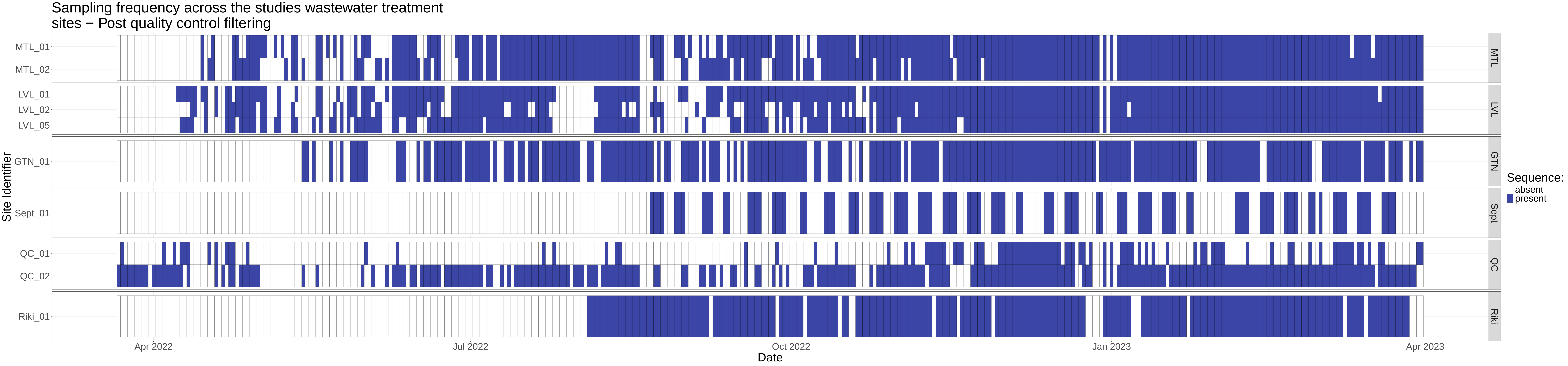

### supp_figure_S5.pdf

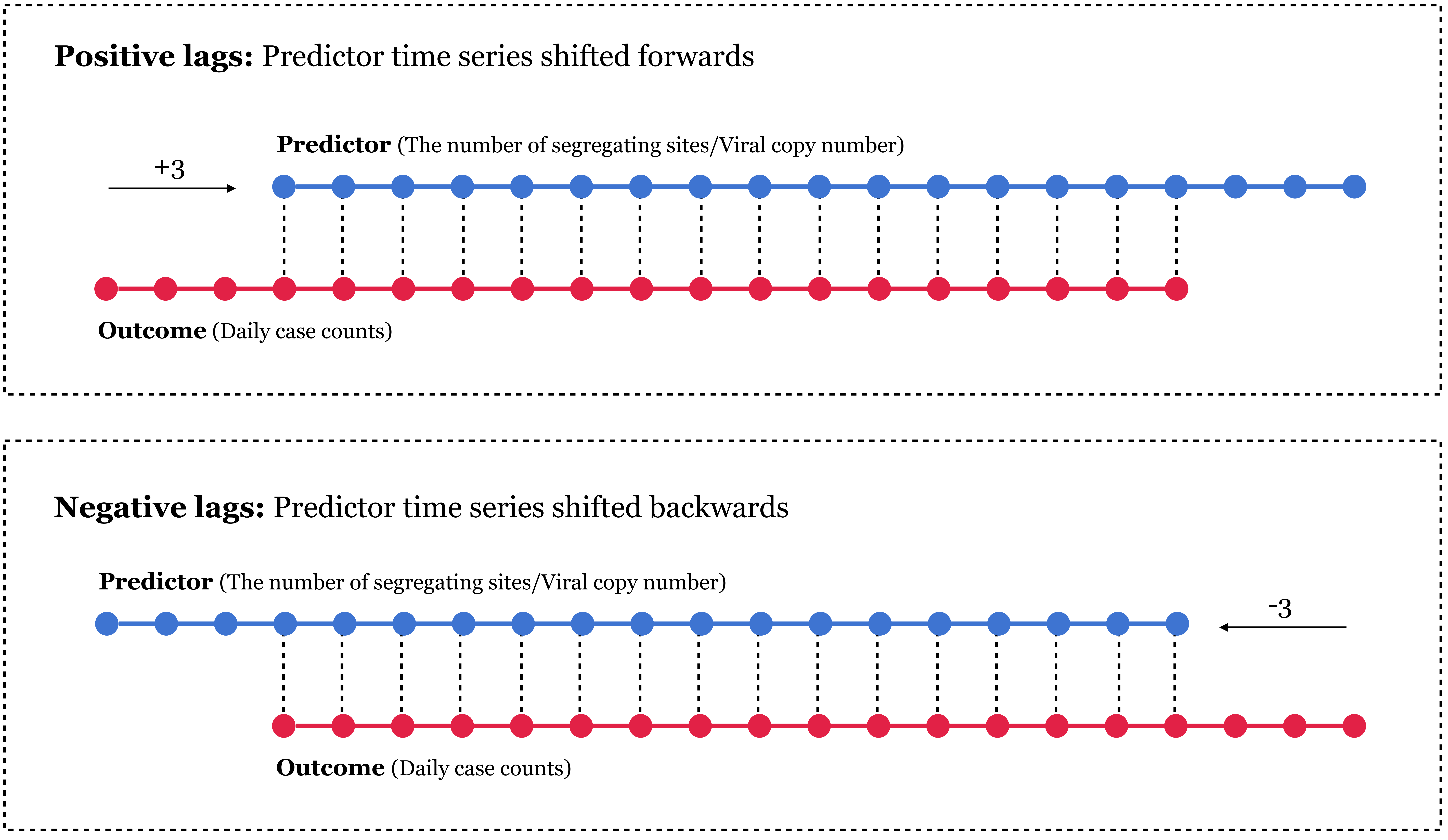

### supp_figure_S6.pdf

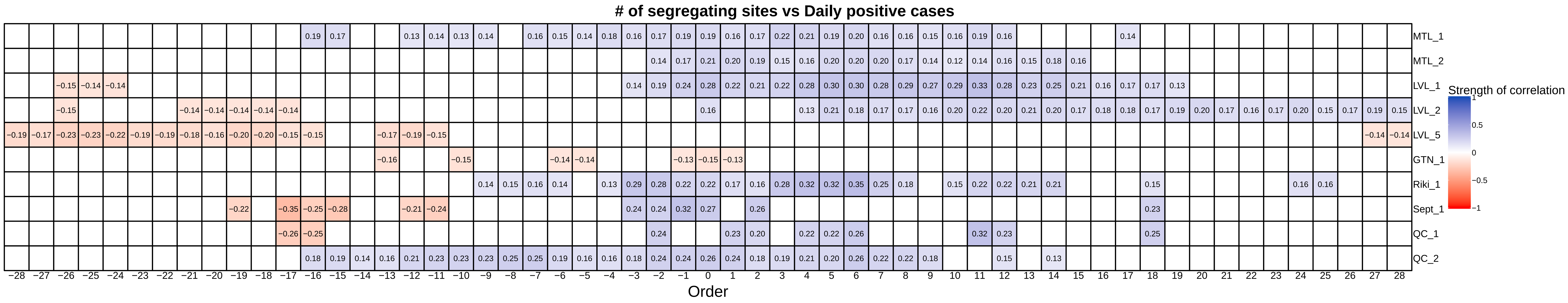

### supp_figure_S7.pdf

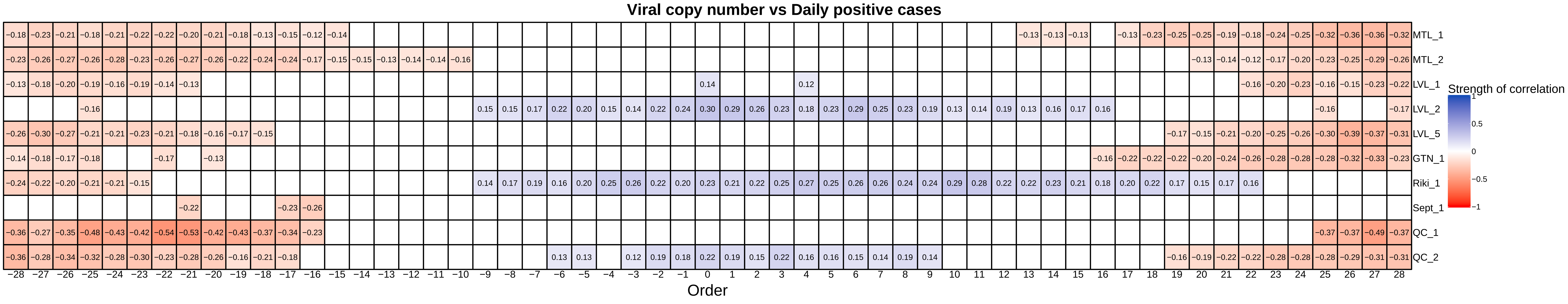

### supp_figure_S8.pdf

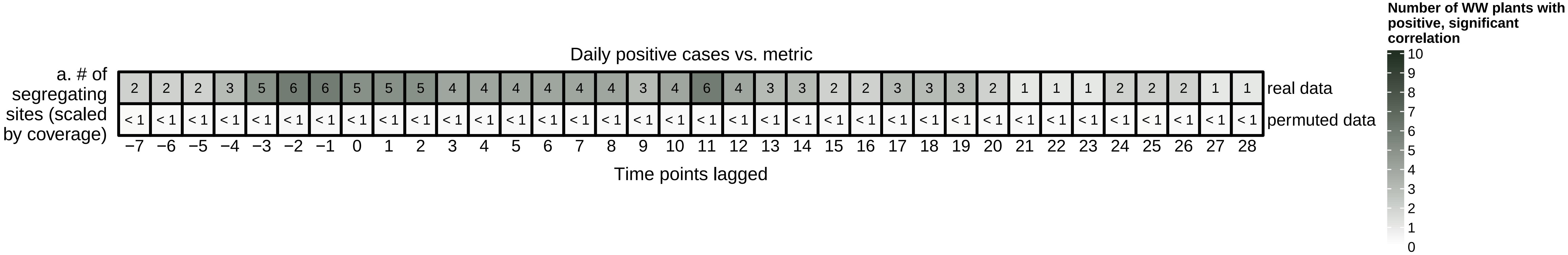

### supp_figure_S9.pdf

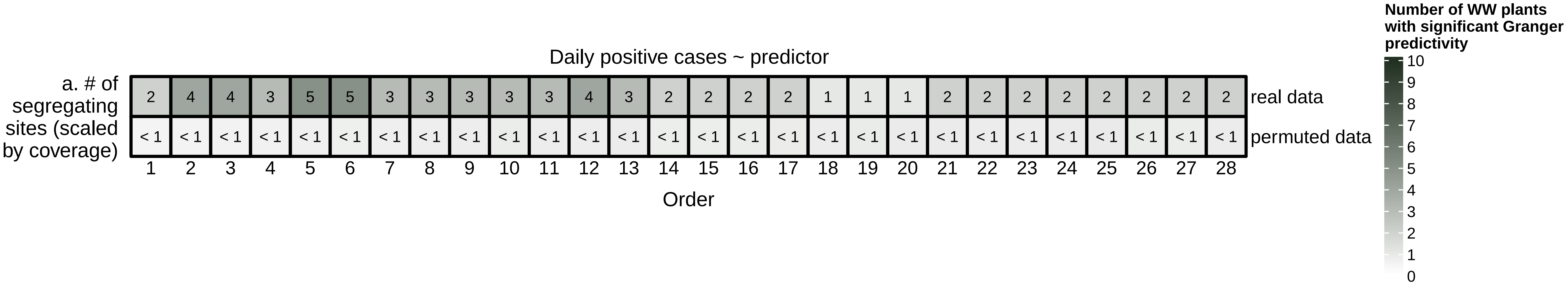

### supp_figure_S10.pdf

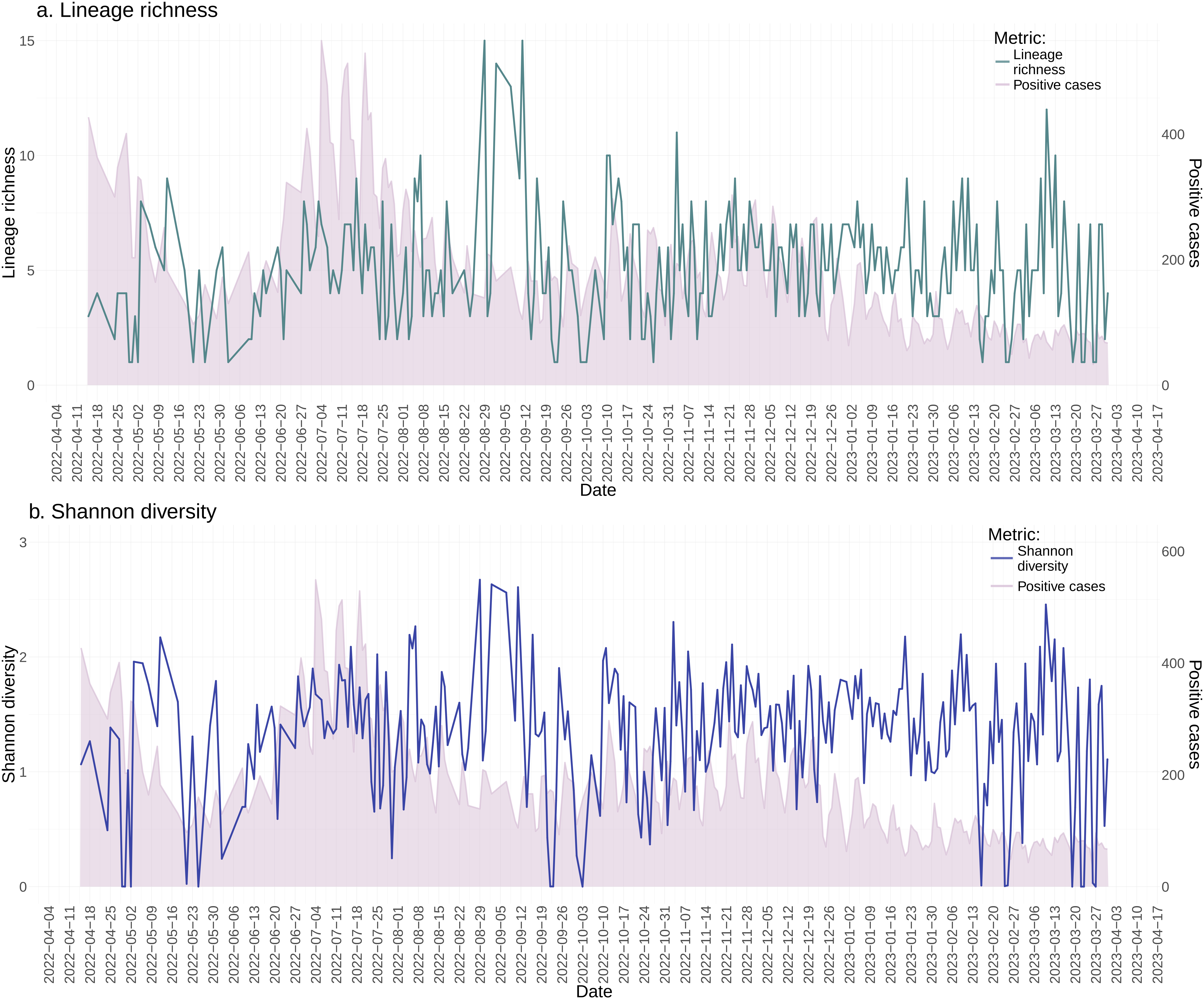

### supp_figure_S11.pdf

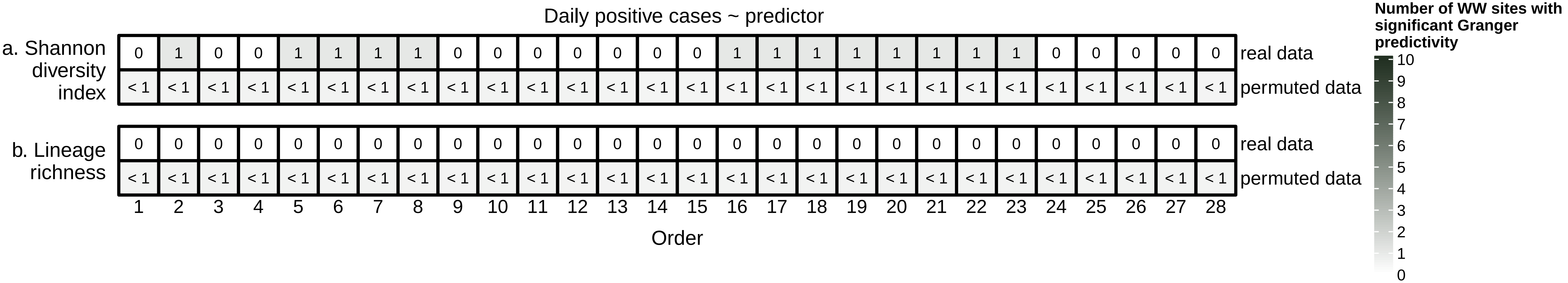

### supp_figure_S12.pdf

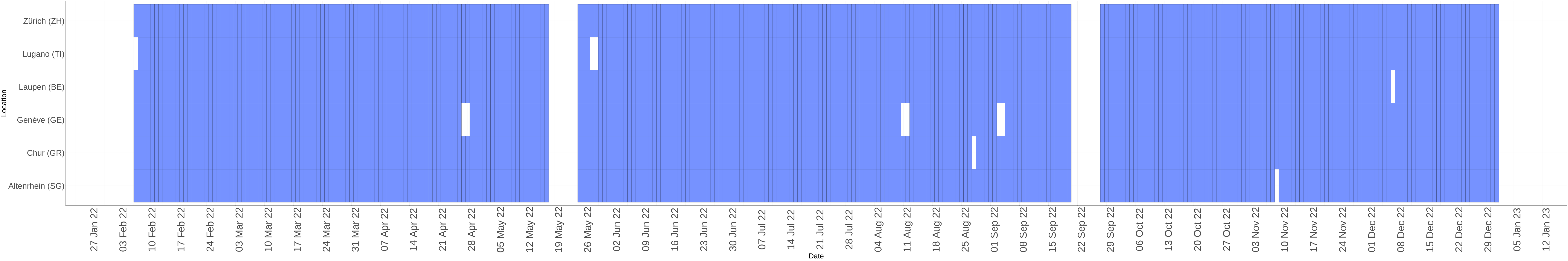
