## Supplementary Methods and Figures for "Predicting COVID-19 case counts using SARS-CoV-2 genetic diversity from wastewater"

**Sample processing and sequencing.** Wastewater samples of 50 mL were concentrated by filtration prior to extraction. Prior to filtration, the initial pH was recorded and adjusted to a final pH of 3.5-4.5 (except for Rimouski samples) and $MgCl_2$ was added to a final concentration of 25 mM. Samples were vacuum-filtered through 0.22 µm (Rimouski) and 0.45 µm (others) pore size diameter mixed cellulose ester membrane filters. Samples were filtered in duplicates, where one filter was archived at -80°C and the other was cut into eight equal pieces and placed into a 2-mL tube for RNA extraction. Samples from Quebec City, Montreal, Laval and Gatineau were processed at McGill University and the samples from Rimouski and Sept-Îles were processed at Université du Québec à Rimouski (UQAR) following the same protocols. Filters were processed using Allprep® Powerviral DNA/RNA kit (QIAGEN) according to the manufacturer's instructions with some modifications. At the lysis step, 10% (v/v) beta-mercaptoethanol and 10% (v/v) bovine rhinotracheitis-parainfluenza$_3$-respiratory syncytial virus vaccine (BRSV, Inforce 3, Zoetis Inc, QC, Canada) were added to the PM1 lysis buffer. Lysis incubation was performed at 55°C for 30 min while rotating. RNA was eluted in 50 µl of RNase-free water and stored at −80°C. The protocol was modified to increase the elution volume to 100 µl starting from 25 October 2022.

Viral genome quantifications for SARS-CoV-2, PMMoV and BRSV were performed in a multiplex RT-qPCR reaction using TaqMan Fast Virus 1-Step Master Mix or Luna® Universal Probe One-Step qPCR and a CFX96 Deep Well™ (McGill University) or Opus 96 Real-Time instrument (UQAR). Technical replicates were prepared for each sample. Thermocycling conditions were performed according to the manufacturer's instructions.

SARS-CoV-2 whole genome amplicon sequencing was conducted using the ARTIC V4.1 protocol (Farr et al., 2022). In brief, RNA extracts were processed for reverse transcription and followed by targeted SARS-CoV-2 amplification using the ARTIC V4.1 primer scheme (https://github.com/artic-network/primer-schemes/tree/master/nCoV-2019/V4.1). Samples were purified and Nextera DNA Flex library preparation was performed for Illumina PE150 paired-end amplicon sequencing on a NovaSeq 6000 instrument at the McGill Genome Centre. Three processing controls were included at the reverse transcription step for quality control of the library preparation workflow and bioinformatic pipeline: NegCtrl_RT (non-template control, as a negative control introduced at reverse transcription step), PosCtrl_LSPQ (RNA of a known viral lineage B.1 isolate from LSPQ as a positive control), and PosCtrl_AccuRNA (synthetic SARS-CoV-2 reference RNA as an additional positive control).

**Sequence processing and lineage calling.** Paired amplicon sequence reads were processed through a wastewater analysis pipeline (see https://github.com/sgsutcliffe/Wastewater_Analysis_Pipeline). First, we removed low-quality reads and reads that mapped to a human reference genome (GRCh38). Reads aligning to the SARS-CoV-2 Wuhan reference genome (MN908947, Wu et al., 2020) were cleaned (primer removal) with iVar 1.3.1 (Grubaugh et al., 2019). We next filtered samples using permissive cutoffs, requiring 10% of the genome to be covered by at least one read, a minimum 100,000 reads total, less than 20% of reads aligning to the human genome, minimum mean depth of 10 reads, and >1% of reads aligning to the reference genome, retaining 2,574 out of 2,747 samples for further consideration in the lineage-based analysis outlined below. As described in the subsequent sections, stricter sequencing depth and variant frequency cutoffs were applied for the calculation of the number of segregating sites. Variant calling, lineage prediction, and relative abundance estimation were performed using Freyja v1.3.12 (Karthikeyan et al., 2022), and lineage calls were binned such that low relative abundance lineages (< 1%) were binned with their parent lineage.

**Swiss wastewater: sample collection and sequencing.** The methodology used for the sample collection and sequencing of the Swiss dataset is previously described (Dreifuss et al., 2025; Jahn et al., 2022; John et al., 2024). Summarizing the work done previously, volume-proportional 24-h composite influent samples were collected, stored at 4 °C for up to five days, and transported on ice to a central laboratory. The samples were processed at a volume of 40 mL using the Wizard Enviro Total Nucleic Acid Extraction Kit from Promega (CN A2991), with an elution volume of 80 ul. Inhibitor removal was performed with OneStep PCR Inhibitor Removal columns from Zymo (CN D6030). RNA extracts were stored at −80 °C for up to 2 weeks. Reverse transcription of the extracted RNA was performed with the LunaScript RT SuperMix kit from New England Biolabs. Tiling amplicon amplification of the cDNA was performed using the ARTIC v4 or v4.1 protocol. Sequencing of the PCR products was done using the Illumina NovaSeq 6000, MiSeq, or Aviti platform, resulting in paired-end reads of 250 bp length each. To only include wastewater sampling locations with sufficiently long time series (minimum of 100 sequences available), we restricted our analysis to samples from Lugano, Werdhölzli (Zurich), Aire (Geneva), Chur, Altenrhein, and Laupen.

**Swiss wastewater: sequence processing and basecount variant files.**
Swiss wastewater sequences were already processed and basecount variant files were available for analysis. Processing of Swiss wastewater sequencing samples and mutation calling had been performed using V-pipe 3.0 (Fuhrmann et al., 2024). In short, V-pipe filtered and trimmed the sequencing reads using PRINSEQ (Schmieder and Edwards, 2011) and aligned them to the SARS-CoV-2 reference genome (GenBank accession number: NC_045512.2) using BWA-MEM (Li and Durbin, 2009). Custom routines implemented within V-pipe were used to generate base counts for each alignment column of each sample (https://www.google.com/url?q=https://github.com/cbg-ethz/smallgenomeutilities/?tab%3Dreadme-ov-file%23aln2basecnt&sa=D&source=docs&ust=1759013146350467&usg=AOvVaw2zI5jC-rhLXLjO-yT9-xDb). The accession lists for the raw wastewater sequence data from Switzerland is included in **Supplementary Table S3**.

**Swiss wastewater: viral copy and positive case count data.** Positive case counts and viral load data were downloaded from the publicly available datasets provided by the Swiss Federal Office of Public Health (FOPH). Positive case counts and viral load data were matched to the basecount files by location and date. An interactive map of the locations of these wastewater sampling locations in Switzerland is available on the dashboard of FOPH (https://www.covid19.admin.ch/en/epidemiologic/waste-water?geoView=table&epiRelDev=abs&geo=CH&rel=abs&wasteWaterFacility=270101).

**Swiss wastewater: Calculating the number of segregating sites from basecount files.** The wastewater sequencing data format readily available were basecount files. Each basecount file corresponding to a wastewater sample, lists the number of A, T, C, G, and "-" genotypes observed at every position along the SARS-CoV-2 genome. Similar to the analysis on the Quebec dataset, for each sample, we excluded genomic positions with less than 50x coverage from the analysis, to ensure enough coverage to be able to detect the number of segregating sites. To determine whether the polymorphism observed at each genomic position is statistically significant, we performed a one-sided binomial test on the two most commonly observed genotypes at that position. Assuming a fixed error rate of 0.3% (Stoler and Nekrutenko, 2021), the number of successes in the binomial test was set to the second most common genotype, and the total number of trials was set to the sum of the two most common genotypes (the assumption being that the *derived* mutation would be the less common genotype of the two). The test was performed with an alternative hypothesis of "greater", and significance was tested at a 0.05 level. The binomial test was carried out in R using the *binom.test*() function, in the *stats* package (R Core Team., 2023). The total number of genomic positions for which the described binomial test was significant was recorded as the number of segregating sites for a given sample.

**Supplementary tables:**

| City ID | sampling location Number | Mean number of segregating sites | Mean number of segregating sites with sample-specific lineage defining mutations removed | Proportion of segregating sites that are lineage defining mutations |
|---|---|---|---|---|
| MTL | 1 | 354.91519434629 | 343.600706713781 | 0.0318794117939885 |
| MTL | 2 | 319.686832740214 | 310.615658362989 | 0.0283751892421408 |
| LVL | 1 | 230.06462585034 | 221.710884353742 | 0.0363104126317657 |
| LVL | 2 | 356.362595419847 | 344.118320610687 | 0.0343590347767412 |
| LVL | 5 | 344.509157509157 | 333.516483516484 | 0.0319082200082934 |
| GTN | 1 | 226.524 | 219.752 | 0.0298952870336035 |
| QC | 1 | 133.474747474747 | 127.373737373737 | 0.0457090964128955 |
| QC | 2 | 235.890977443609 | 227.093984962406 | 0.0372926195674694 |
| Riki | 1 | 383.022522522523 | 365.54954954955 | 0.0456186567251943 |
| Sept | 1 | 206.12380952381 | 196.171428571429 | 0.048283509679804 |

**Table S1.** The average number of segregating sites found at every sampling location, before and after removing the lineage defining mutations. The difference in percentages is shown.

**Table S2**: Table of the GISAID IDs for the clinical sequences used in this analysis, and their corresponding lineage calls. Subjects for which no GISAID ID was available are marked as "unavailable".

**Table S3.** The metadata and ENA accession IDs for the raw wastewater sequence data from Switzerland

**Supplementary figures:**

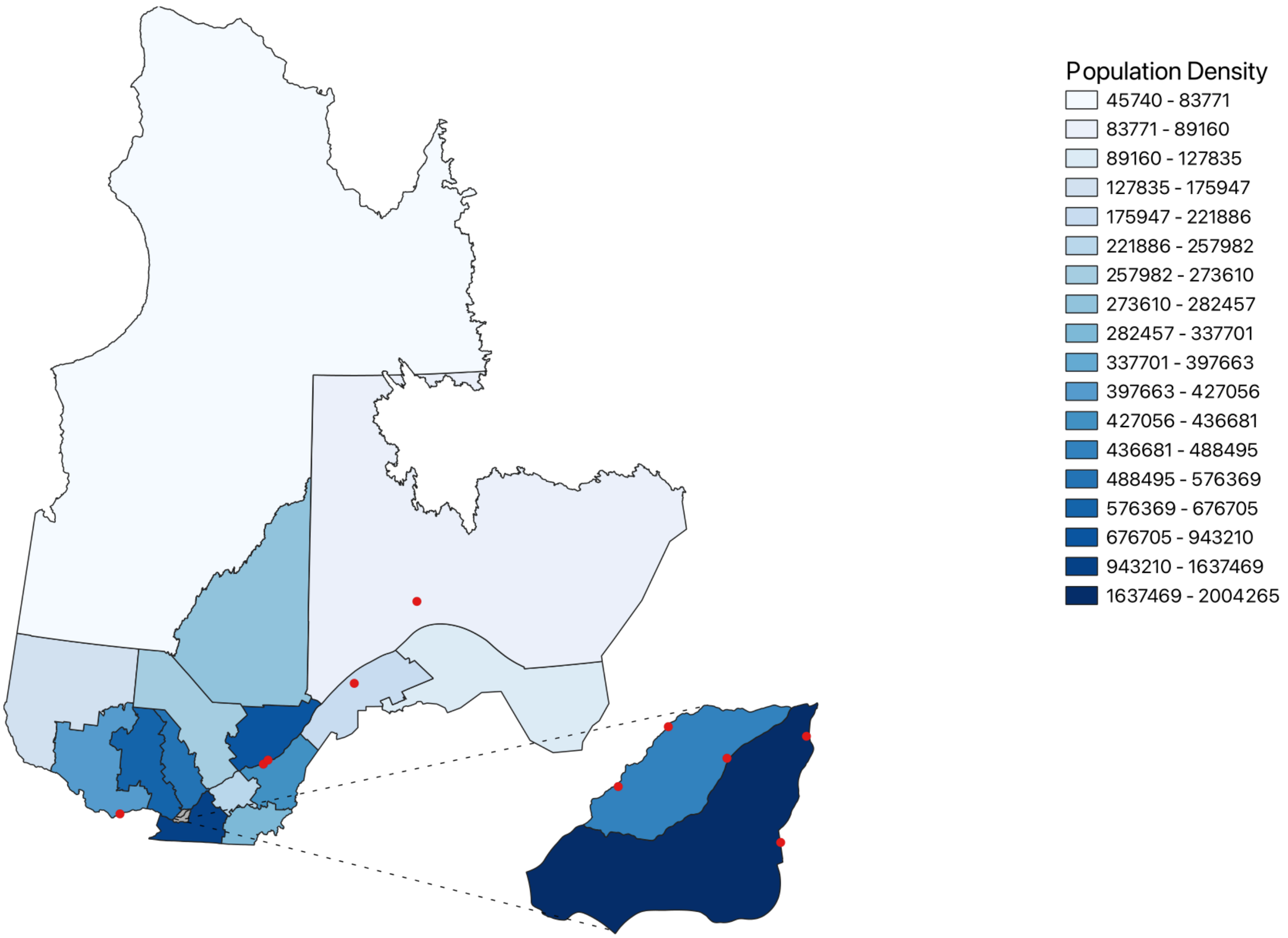


**Supplementary Figure S1. Map of the province of Quebec, displaying population density and wastewater sampling locations studied.** The plot shows population density in shades of blue with sampled wastewater locations marked with red points. The cities of Laval (top, lighter blue) and Montreal (bottom, dark blue) are shown in the zoom region at the bottom right (not to scale).

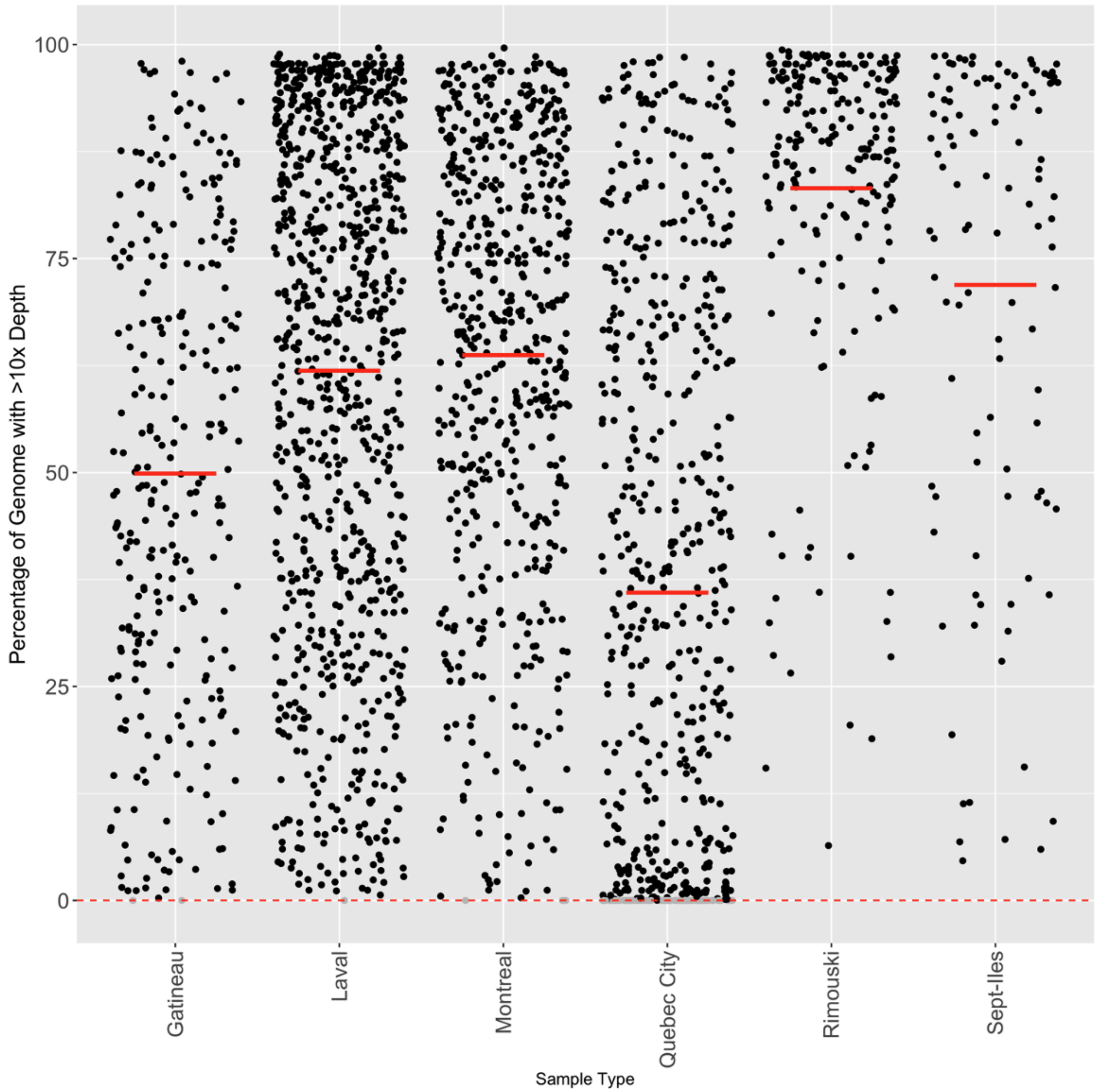


**Supplementary Figure S2. Wastewater sample quality by city.** Quality of samples prior to removal of low-quality samples. Sample quality per sample calculated by Freyja (percentage of genome breadth with a read-depth greater than 10x). *Red bar*: Mean coverage. *Dotted line*: 0.01 coverage.

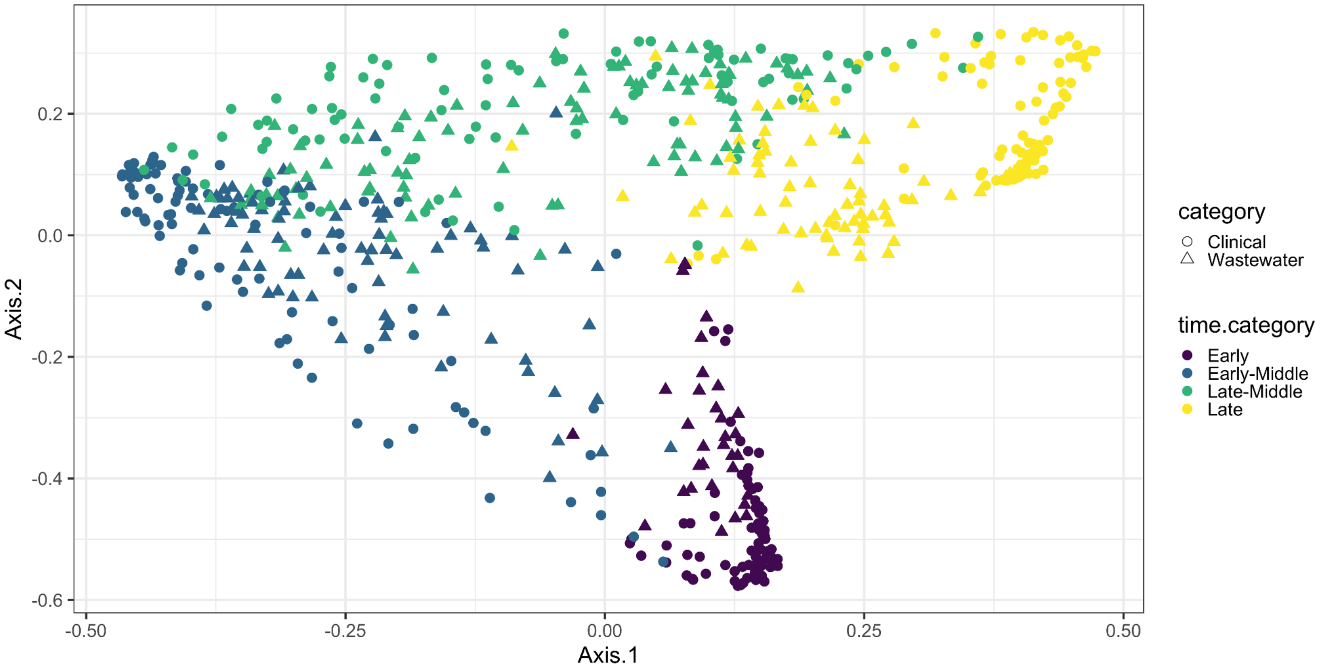


**Supplementary figure S3. SARS-CoV-2 lineage composition varies more over time than by sample type (clinical or wastewater).** The first two axes are shown from a PCoA on Bray-Curtis distances among SARS-CoV-2 lineages found in either wastewater or clinical samples. Samples were categorized temporally by Early, Early-Middle, Late-Middle, and Late based on a 16-week interval of sampling collection. The colors of data points indicate the temporal categorization of samples, and shapes indicate sample type (clinical, wastewater). Samples are more closely clustered by temporal label than they are by sample type.

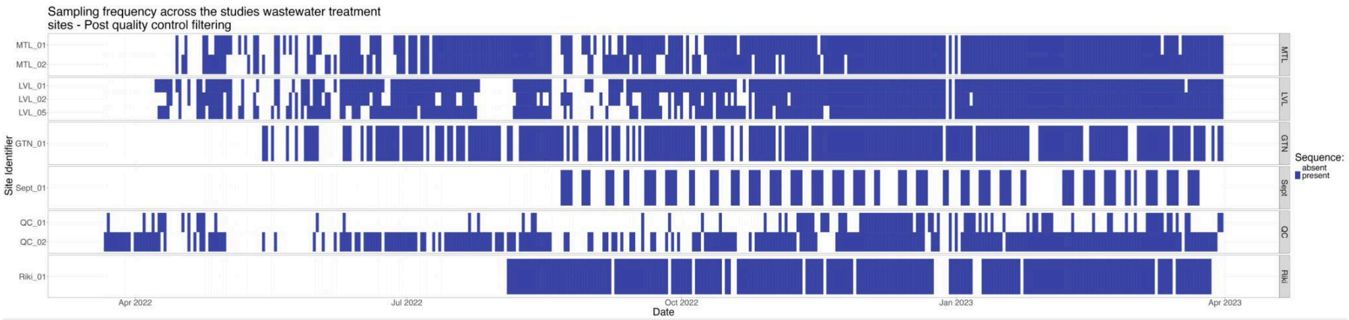


**Supplementary figure S4. Sampling frequency of time series used across wastewater sampling locations in Quebec, post quality control filtering of sequences.**

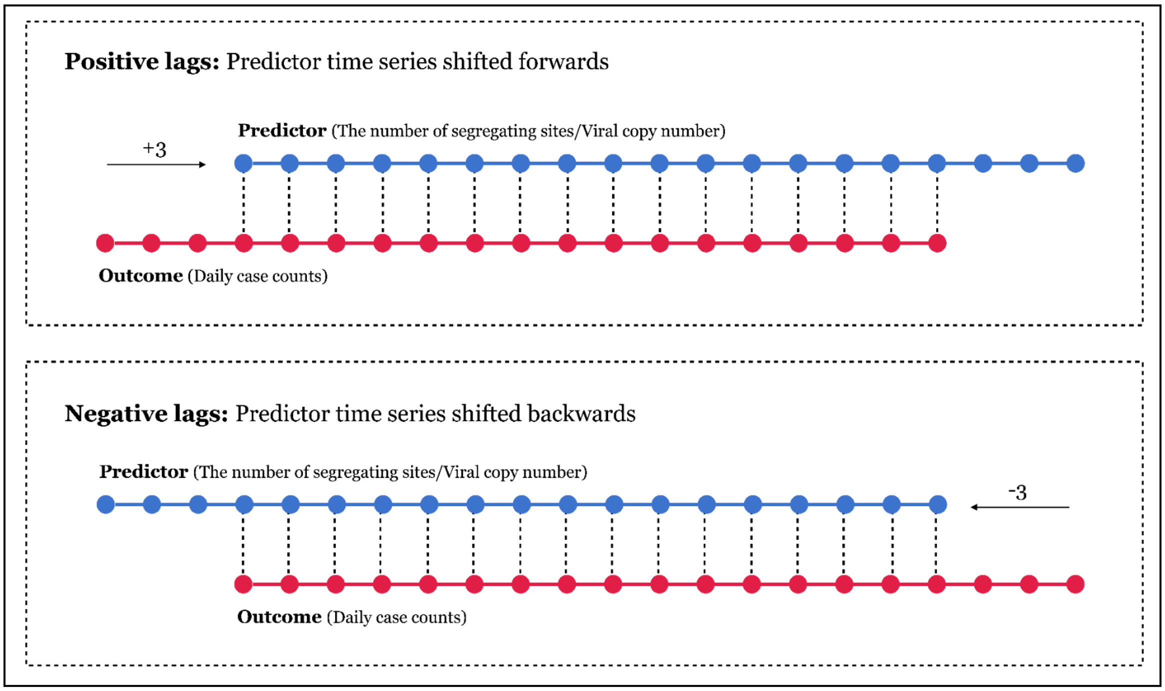


**Supplementary figure S5. Schematic definition of lags for the Spearman's correlation test.** Positive lags mean that the predictor time series was shifted forward, resulting in earlier values of the predictor time series to be paired with later values of the outcome time series when testing for a monotonic relationship in the Spearman's correlation test. Negative lags mean that the predictor time series was shifted backwards so that later values of the predictor time series were paired with earlier values of the outcome time series in the Spearman's test.

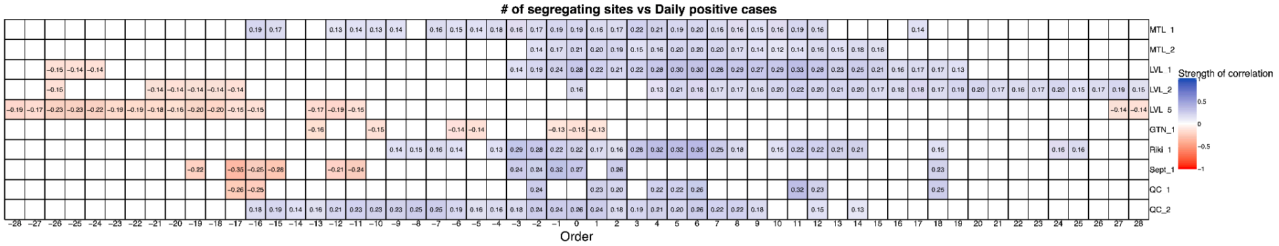

**Supplementary figure S6. Grid of Spearman's correlation values between the number of segregating sites and positive case counts.** The x axis shows the lag applied for the test, and the y axis shows wastewater sampling locations. The numbers in the cells show the estimated correlation coefficient for significant tests. Spearman's correlations not significantly different from zero are shown in white, and significant tests are shown in shades of blue (positive correlation coefficients) and red (negative correlation coefficients). Significance was tested at a 0.05 level.

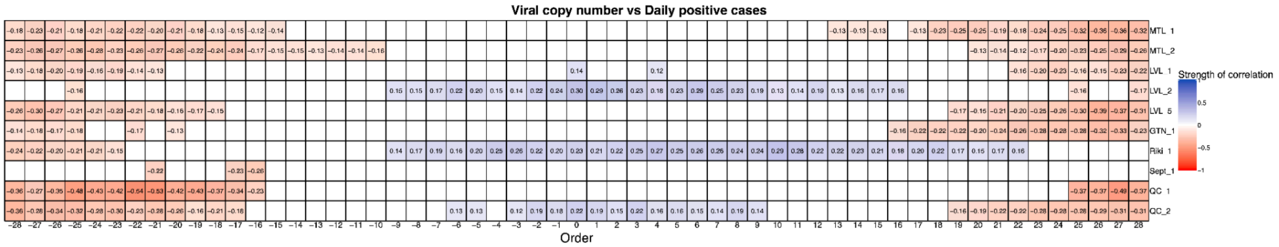

**Supplementary figure S7. Grid of Spearman's correlation values between viral copy number and positive case counts.** The x axis shows the lag applied for the test, and the y axis shows wastewater sampling locations. The numbers in the cells show the estimated correlation coefficient for significant tests. Spearman's correlations not significantly different from zero are shown in white, and significant tests are shown in shades of blue (positive correlation coefficients) and red (negative correlation coefficients). Significance was tested at a 0.05 level.

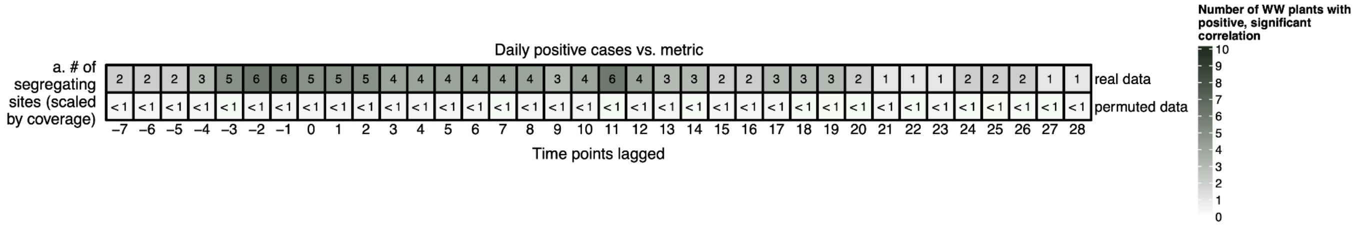


**Supplementary figure S8**. **Summary of Spearman's correlation test between the number of segregating sites per sufficiently covered genomic position, with positive case counts**. The top row shows the number of wastewater sampling locations in which a significant, positive Spearman's correlation between the number of segregating sites per sufficiently covered genomic position, and positive case counts was observed. The bottom row shows the proportion of "false-positive" tests obtained on permuted data. Lags -7 to +28 are shown, and the direction of the lags are schematically described in Supplementary figure S1.

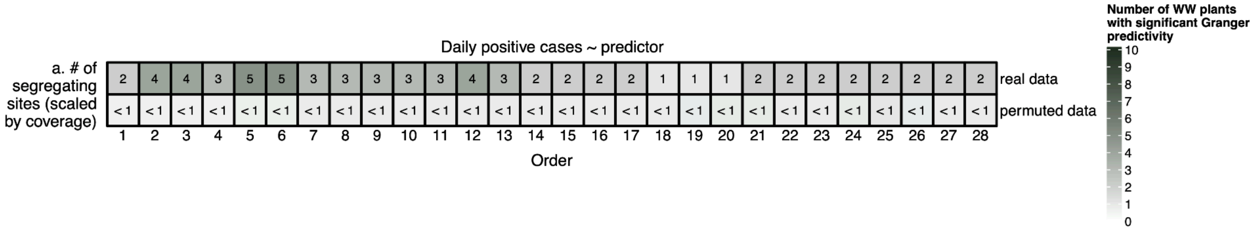


**Supplementary figure S9. Summary of Granger test for the number of segregating sites per sufficiently covered genomic position, predicting positive case counts.** The top row shows the number of wastewater sampling locations in which a significant Granger relationship exists. The bottom row shows the proportion of "false-positive" tests obtained on permuted data.

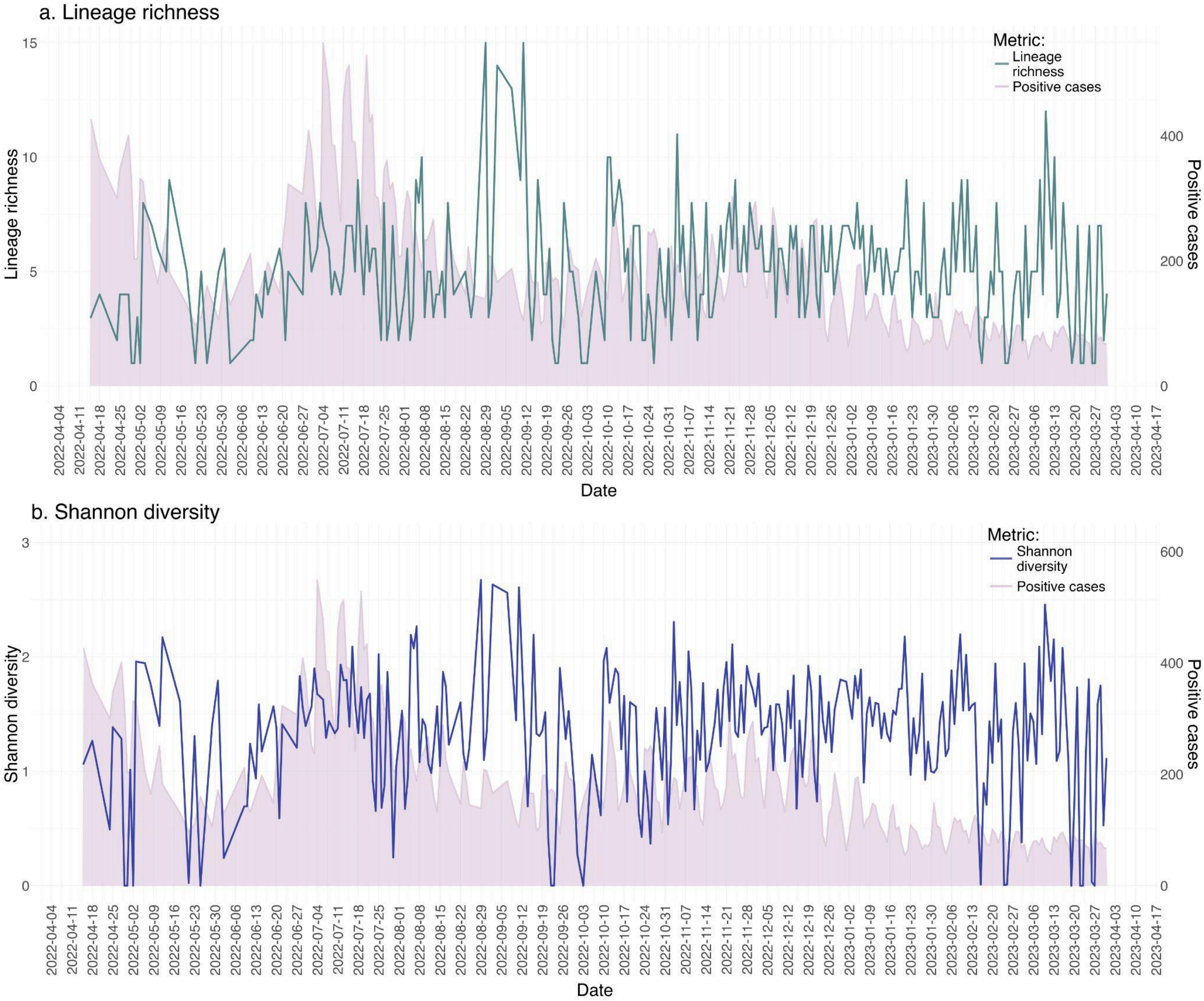


**Supplementary figure S10. Shannon diversity and lineage richness in Montreal sampling location 1.** Time series of lineage richness (top) and Shannon diversity (bottom) in one of the two wastewater sampling locations in Montreal.

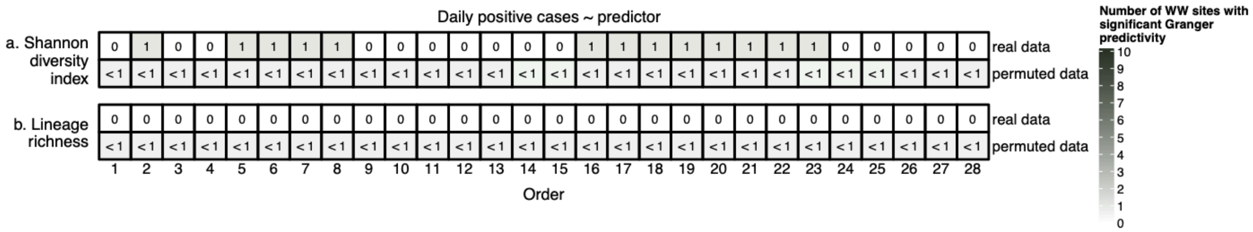


**Supplementary figure S11. Summary of Granger test for lineage metrics predicting positive case counts.** Shannon diversity index (a) and lineage richness (b) perform poorly in predicting daily case counts.

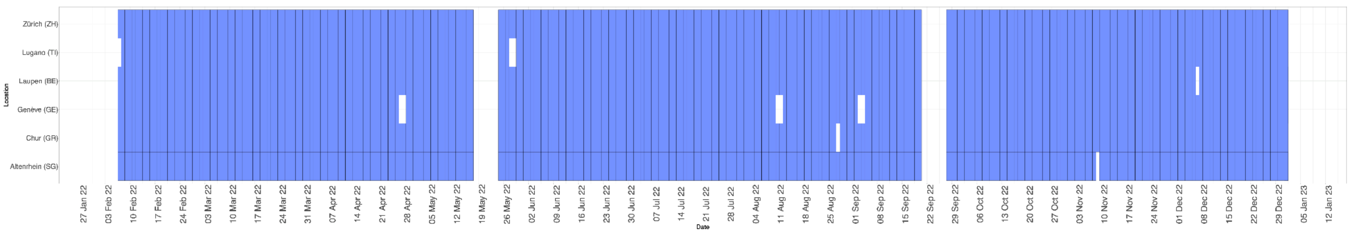


**Supplementary figure S12.** Sampling frequency of time series used across wastewater sampling locations in Switzerland. The distribution of the sequencing used in the time series analysis after matching to available case counts and viral load data are shown.
